# Retinal Thickness in Anxiety, Depression, and Substance Use Disorders: A Systematic Review and Meta-Analysis of Optical Coherence Tomography (OCT) Studies

**DOI:** 10.64898/2026.03.19.26348797

**Authors:** Michaela Joan Grimbly, Sheri-Michelle Koopowitz, Ruiye Chen, Wenyi Hu, Zihan Sun, Paul J Foster, Dan J. Stein, Zhuoting Zhu, Jonathan Ipser

## Abstract

**Background:** Optical coherence tomography (OCT) is increasingly used to investigate retinal structural changes across neurological and neuropsychiatric conditions. This systematic review and meta-analysis synthesises evidence examining retinal thickness in anxiety, depression, and substance use disorders (SUD) compared with healthy controls.

**Methods:** A pre-registered systematic search (PROSPERO: CRD42024559542) of major databases following PRISMA guidelines was conducted. Case-control studies measuring retinal layer thickness via OCT in adults with DSM or ICD diagnosed anxiety, depression, or SUD were included. Multilevel random-effects models were used to calculate pooled standardised mean differences (SMD) and account for dependencies.

**Results:** Thirty-three studies were included for narrative review, and 25 studies with 145 effect sizes were included for meta-analysis. The primary analysis, which pooled all disorders and effect sizes from available retinal thickness measures, found no significant differences between cases and controls (SMD = −0.20; 95% CI [−0.53, 0.14]; p = .244). Subgroup analyses for anxiety, depression, and SUD also yielded non-significant results (all p > .05). No specific retinal layer was consistently affected, and there was no evidence of an age × diagnosis interaction. Significant heterogeneity (Q = 756.57, p < .001) was present across analyses.

**Conclusion:** This meta-analysis found no significant associations between retinal structural differences and anxiety, depression, or SUD. The field is characterised by high heterogeneity and publication bias, limiting the strength of evidence for the utility of OCT as a reliable biomarker for these conditions. Standardised, large-scale studies are needed with strict controls for confounding factors, including medication, disease stage and ocular parameters, alongside standardised OCT segmentation protocols.

**Article Highlights:**

- First meta-analysis of OCT retinal thickness in anxiety, depression and SUD.
- No significant retinal thickness differences found between cases and healthy controls.
- Age and sex did not moderate the association between diagnosis and retinal thickness.
- High heterogeneity and publication bias limit utility of OCT as a neuropsychiatric biomarker.
- Standardised protocols are needed to clarify retinal changes in psychiatric research.

## Introduction

### Rationale

Common mental disorders are highly prevalent, with a global lifetime aggregate prevalence estimated at 29% (Steel et al., 2014). Depression and anxiety are the most frequent of these conditions, and in 2021, they accounted for over 63% of the total global burden of all mental disorders(Zhang et al., 2026). Furthermore, mental and substance use disorders collectively contribute to overall health loss, responsible for approximately 20% (17.2% and 2.3% respectively) of years lived with disability (YLDs) globally(World Health Organization, 2025). Despite research advances, the pathophysiological mechanisms underlying these disorders are poorly understood, with complex biological, environmental and genetic factors believed to play a role in their aetiology (Bora et al., 2012; Maes et al., 2018; Malhi and Mann, 2018; Volkow and Boyle, 2018). A significant barrier to effective care is that current mental health diagnostics rely on self-reported symptoms. This is inherently subjective, often resulting in mis- and underdiagnosis of common mental disorders (Vermani et al., 2011). Consequently, there is an urgent need for innovative screening methodologies that can provide more objective and quantifiable measures of mental health status.

One promising candidate for such an objective method is optical coherence tomography (OCT). The retina, a readily accessible neurological structure, serves as a proxy for studying the brain due to its underlying embryological and anatomical similarities to the central nervous system (Foster and Khaw, 2009; London et al., 2013).OCT, a light-based technique that images the retina has already demonstrated significant utility in neurology. It is currently used to monitor neuroaxonal loss in Multiple Sclerosis (Petzold et al., 2021) and serves as a non-invasive proxy for cortical amyloid and tau pathology in Alzheimer’s disease(Cheung et al., 2021). Furthermore, large-scale studies have demonstrated that lower retinal thickness is a predictor of incident dementia (Mutlu et al., 2018), cognitive decline (Ko et al., 2018) and both prevalent and incident Parkinson’s disease(Wagner et al., 2023). The retina likewise may similarly serve as an accessible index of neurobiological changes in psychiatric illness(Gupta et al., 2023).

In addition to neurodegenerative disease, the retina has also been shown to reflect the biological ageing process, typically manifesting as thinning in the retinal nerve fibre layer (RNFL), ganglion cell layer (GCL) and retinal pigment epithelium (RPE) (Bonnel et al., 2003; Demirkaya et al., 2013; Leung et al., 2012; Sung et al., 2009). Beyond these ageing changes, lower retinal thickness has been reported across a spectrum of conditions, including schizophrenia spectrum disorders (Gonzalez-Diaz et al., 2022; Komatsu et al., 2022), major depressive disorder(Xiao et al., 2024), bipolar disorder (Prasannakumar et al., 2023), and neurocognitive impairment (Kim et al., 2022; Noah et al., 2020). The theoretical rationale for using the retina as a proxy for the brain rests on their shared embryological and anatomical ties, whereby the retina, an extension of the central nervous system (CNS), often mirrors CNS health(London et al., 2013). As common mental disorders are often associated with amplified age-related changes in the brain, such as volume atrophy and neuronal loss, (Kaufmann et al., 2019; Wrigglesworth et al., 2021), it stands to reason that OCT has potential as a biomarker for neuropsychiatric disease.

Existing literature in this field is, however, characterised by significant inconsistency, with individual studies reporting conflicting findings regarding retinal structural differences. While retinal thinning has been established in psychiatric conditions like schizophrenia, the evidence for other common and often comorbid conditions remains relatively sparse and layer-specific. For instance, while lower GCIPL thickness has been reported in major depressive disorder across several studies, findings for the RNFL remain highly divergent, with some reports showing significant thinning and others no identifiable differences. These inconsistencies likely stem from high clinical and methodological heterogeneity, including confounding factors such as medication use, comorbidities or illness duration.

Investigating retinal structural changes in these conditions, while accounting for age-related changes, is an important avenue to pursue, especially given the frequent overlap in the occurrence of common mental disorders, such as anxiety, depression, and substance use. Since these disorders have all been associated with accentuated biological ageing (Gao et al., 2023; Kallen et al., 2024; Scheffler et al., 2024; Zhu et al., 2026), it is also important to determine if the relationship between age and retinal thickness is amplified in cases compared to controls.

This systematic review and meta-analysis aims to provide a comprehensive review of the existing literature on retinal thickness measures in anxiety, depression and SUD, as compared to healthy controls. It aims to review the OCT literature to answer the following questions:

1. Are there differences in the retina in those individuals diagnosed with anxiety, depression, or SUD, relative to healthy controls?
2. Are retinal differences consistent across disorders? Are there specific retinal layers that are more affected by diagnoses?
3. Is there an age × diagnosis interaction, whereby the association between ageing and retinal thickness measures is amplified in cases compared to controls?

## Methods

We followed the Preferred Reporting Items for Systematic Reviews and Meta-Analyses (PRISMA) guidelines. The review protocol is registered to the International Prospective Register of Systematic Reviews (PROSPERO) with registration number: CRD42024559542.

### Search strategy

A systematic literature search was conducted from 7 - 8 July 2024 in Pubmed, Scopus, Web of Science, MedRxiv, BioRxiv and ClinicalTrials.gov to identify relevant articles published since the inception of these databases. Initial search terms for the Pubmed database included the MESH terms, (“;Tomography, Optical Coherence”[Mesh]) AND “;Substance-Related Disorders”[Mesh]; (“;Tomography, Optical Coherence”[Mesh]) AND (“;Anxiety”[Mesh] OR “;Anxiety Disorders”[Mesh]); (“;Tomography, Optical Coherence”[Mesh]) AND (“;Depression”[Mesh] OR “;Depressive Disorder”[Mesh]). The search was run individually for each disorder of interest and amended for each database searched. There were no further filters set or date or language restrictions applied. To ensure currency, the search was updated on Pubmed on 12 January 2026. This updated search identified three additional articles which were included for narrative review. Refer to the Supplementary Table S1 for the full search strategy used.

### Eligibility criteria

Published studies of individuals diagnosed with anxiety disorders, major depressive disorder and/or substance use disorder, regardless of treatment status or duration of abstinence, using instruments based on the DSM III+, or ICD 9+ criteria were included for analysis. Studies were restricted to adult humans, but no language criteria were imposed. Posterior segment optical coherence tomography, a non-invasive, light-based imaging technique, was used to assess retinal structures (Zeppieri et al., 2023). Studies with a primary focus on OCT angiography (OCTA) were included only when structural thickness measures were derived using segmentation algorithms equivalent to conventional OCT protocols. Studies that did not mention DSM or ICD diagnostic criteria were excluded from comparative analysis.

### Selection process

The initial search retrieved 958 publications across all databases that were potentially eligible. Following deduplication (n = 660), study titles and abstracts were independently screened by two reviewers (MG, SK) according to eligibility criteria. If deemed potentially eligible by at least 1 reviewer, the corresponding full-text article was screened by the same reviewers (n = 77). A third reviewer (JI) resolved conflicts. Five full-text articles were not retrievable and therefore were excluded. The 30 studies from the initial search that passed the full-text screen were then subjected to critical appraisal by the 2 individual reviewers (MG, SK) using the JBI Critical Appraisal Tool for Case-Control Studies (Moola S, 2017). This procedure was also employed for the three additional studies identified during the updated search. Reference lists of the articles selected for inclusion were screened, and potentially eligible articles were subjected to the same process for inclusion. A total of 33 articles were included for narrative review, of which 25 were included for meta-analysis. Please see Figure 1: PRISMA Diagram.

**Figure 1:**
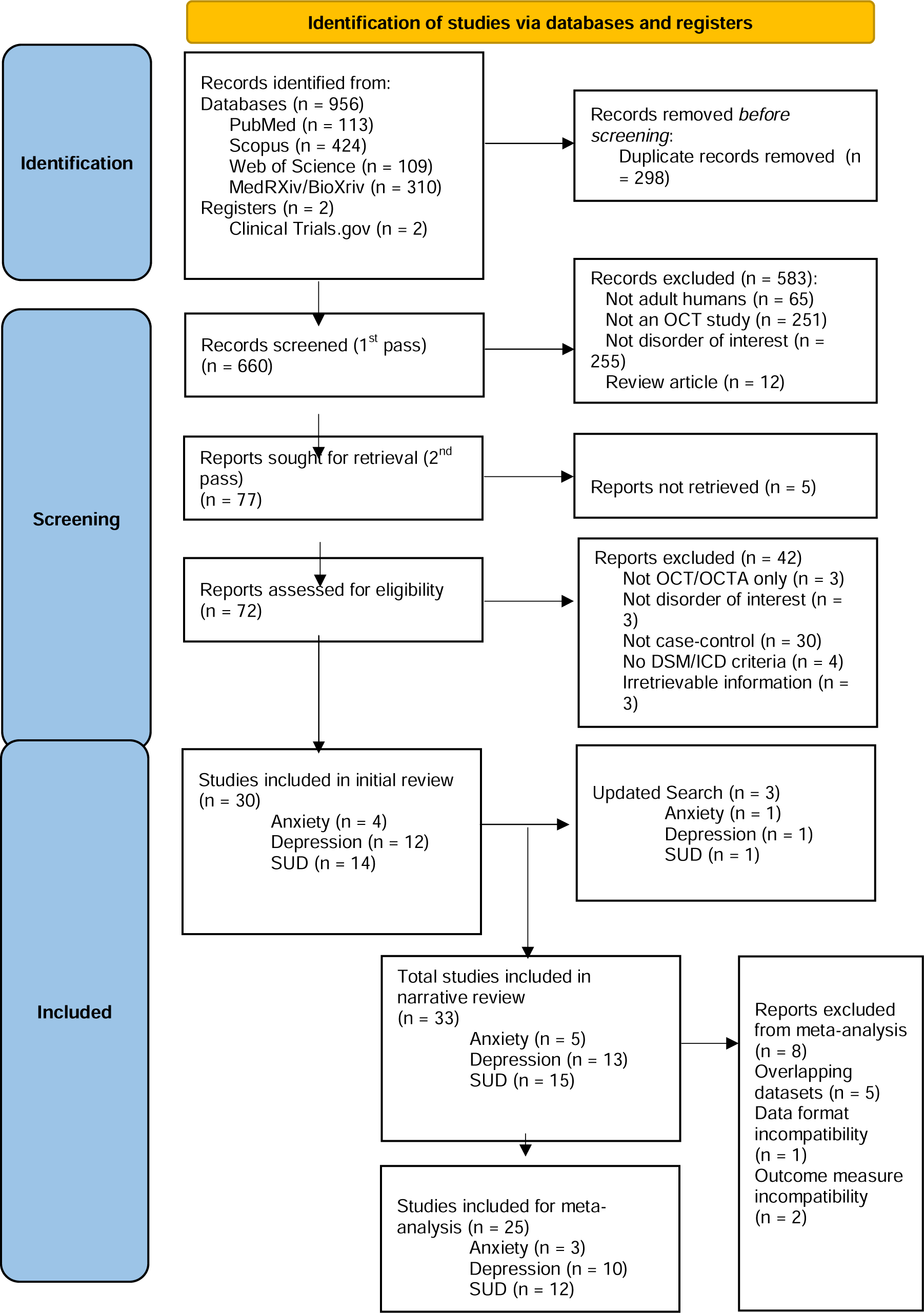
PRISMA Diagram

### Data collection process

The following data were extracted into a customised spreadsheet: study site, recruitment timeline, publication date, primary diagnosis, exclusion criteria, treatment, diagnostic criteria used, questionnaires used, OCT model used, eye imaged, relevant measurements for each retinal layer report, study sample characteristics, including age and sex distribution, main outcomes and secondary findings. In cases where data was missing, the corresponding authors were contacted for additional information. Thirty-three articles were included for narrative review following the screening process, as they met all eligibility criteria. A subset of 25 studies were included for meta-analysis, on the condition that they provided data that could be used for group comparisons, such as means and standard deviations. For the meta-analysis, we aimed to synthesise evidence on retinal thickness layers across individuals with anxiety, depression, and substance use compared to healthy controls. Where samples overlapped between studies, the study with the largest sample was included. Meta-analyses were conducted only for outcomes reported in 3 or more studies.

### Statistical Analysis

Statistical analyses were conducted using R (version 4.5.2) with the *metafor* package (version 4.8-0) (Viechtbauer, 2010). The Restricted Maximum Likelihood (REML) method was used for all analyses described below, unless otherwise indicated. Mean thickness measures, standard deviations and sample sizes were extracted from each of the studies included. Effect sizes were calculated as standardised mean differences (SMD) using *escalc* for every available retinal layer measurement, with a confidence interval (CI) of 95%. A p-value of less than 0.05 was considered statistically significant.

To address the dependency issue of multiple effect sizes originating from the same study, a multilevel random effects model was used, employing *metafor rma.mv* (Gooty et al., 2019). For example, this accounted for dependencies where studies reported measurements for both the right and left eye, or across several retinal layers from the same group of participants. A two-level meta-analytic model was run to handle the statistical dependencies arising from multiple measures originating from the same study to prevent overestimation of precision by ensuring each study contributes appropriately to the overall results. Effect sizes were first calculated individually, then aggregated into broader subsets (e.g. all superior RNFL measures, temporal and nasal) for overall meta-analysis of regions.

We analysed studies by first comparing all cases, irrespective of diagnosis to controls, then individually in pairwise comparisons between each diagnostic group (anxiety, depression, and substance use disorder) to controls. Finally, a three-way analysis was conducted to simultaneously compare all three disorders with one another. A sensitivity analysis was run, removing potential outliers defined as effect sizes falling outside of the 95% pseudo-confidence interval limits upon visual inspection of the forest plots. To test for an age × diagnosis interaction, we conducted a meta-regression analysis using mean age as a continuous moderator.

Given the substantial variability in retinal layers reported across studies, the primary analysis intentionally aggregated all available thickness measures to provide a global estimate of retinal structural differences across disorders. This was complemented by subgroup and layer-specific analyses.

### Publication bias

To assess potential publication bias, Egger’s regression tests were run for each of the defined meta-analytic models, alongside visual inspection of the funnel plots for asymmetry. In each regression, the effect size was regressed on its standard error to assess the relationship between study precision and its observed effect size. Between-study heterogeneity was assessed separately using a Q statistic and tau^2^within a random effects framework. A statistically significant Q-test (p < .05) was used as the criterion to indicate the presence of heterogeneity requiring further investigation through moderator analyses.

## Results

A total of 33 studies were included for narrative review and 25 for meta-analysis.

### Study characteristics

A summary of the 33 included studies’ characteristics is illustrated in Table 1. All studies included were case-control studies published between 2016 and 2025. Five studies reported on samples diagnosed with anxiety disorders (Acan et al., 2024; Baykara et al., 2024; Baykara et al., 2022; Kalenderoglu et al., 2025; Ozisik and Kiraz, 2023), thirteen studies of patients diagnosed with depression (ElShaarawi et al., 2022; Friedel et al., 2025; Genc et al., 2019; Jung et al., 2020; Liu et al., 2022a; Liu et al., 2022b; Liu et al., 2021b; Schönfeldt-Lecuona et al., 2018; Sönmez et al., 2017; Turhan et al., 2024; Wang et al., 2024; Xiao et al., 2024; Yildiz et al., 2016) and 15 studies of individuals with substance use disorders (SUD) (Álvarez-Sesmero et al., 2019; Dayi et al., 2020; Demir et al., 2022; Gemelli et al., 2019; Kalenderoglu et al., 2020; Karadere et al., 2020; Kaya and Kaya, 2023; Kaya and Kaya, 2024; Khalil et al., 2023; Liu et al., 2021a; Orum and Kalenderoglu, 2020, 2021; Orum et al., 2022; Özsoy et al., 2023; Talebnejad et al., 2020). Three studies did not mention the machine used for retinal scan acquisition. Thirty-two of the 33 studies included age demographic data. Eight studies were excluded from meta-analysis to maintain statistical homogeneity and independence. (See Figure 1: Prisma flowchart)

**Table 1:**
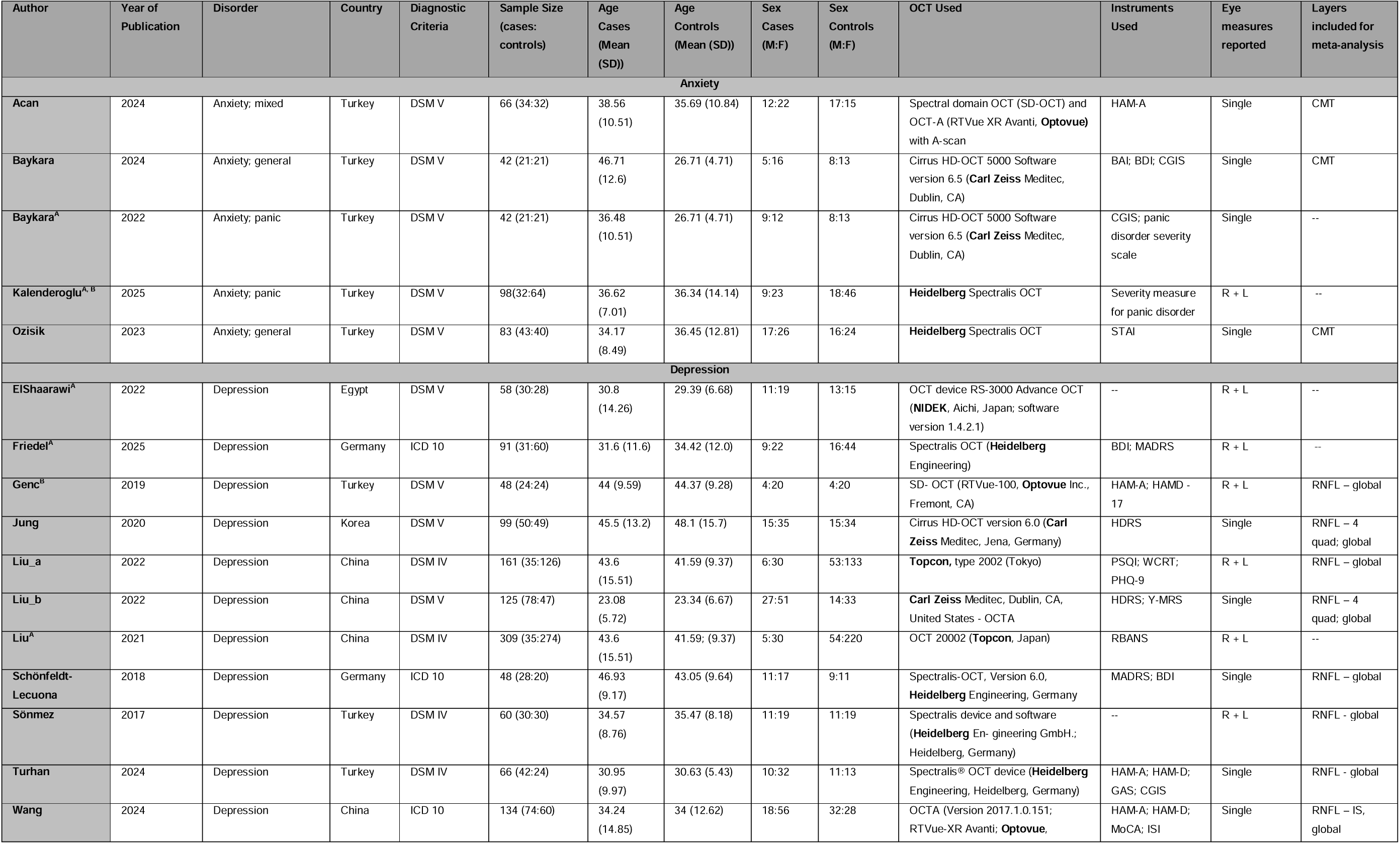
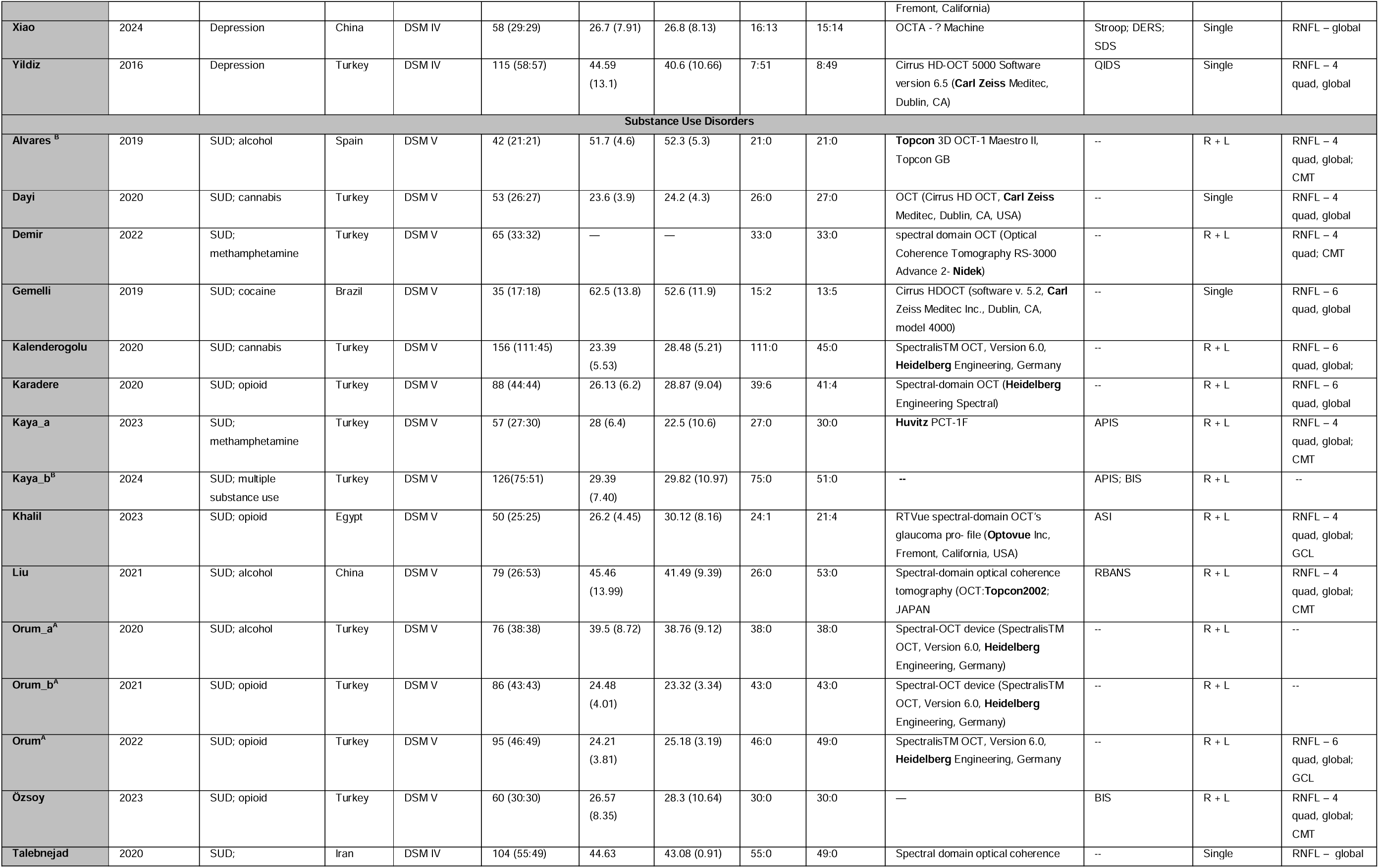
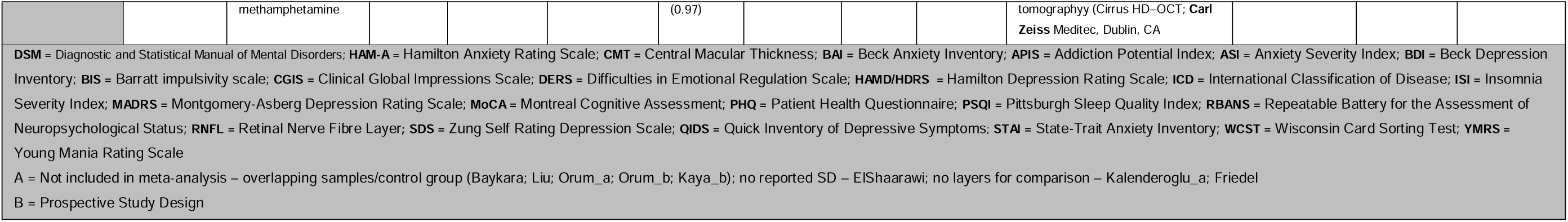
Study Characteristics.

### Risk of Bias Assessment

Overall, the methodological quality of the included studies was variable. The most common limitation was inadequate identification or management of potential confounders noted in 29 of the 33 included studies. These studies failed to report on, or adjust for, potential confounders such as medication status, illness duration, smoking or other ocular parameters. Secondly, group comparability was a weakness across several studies, as statistically significant age differences were reported between cases and controls(Baykara et al., 2024; Baykara et al., 2022). Furthermore, few studies failed to report on recruitment methods or criteria used to define “healthy” controls. Further limitations included small sample sizes (< 50 participants in 7 studies), single-sex samples (12 of the 15 studies included in the SUD sample were male-only), and inconsistent application of diagnostic frameworks and diverse assessment instruments.

### Narrative Review

#### Anxiety

Five case-control studies from Turkey published between 2023 and 2025 examined OCT differences in anxiety disorders (Panic disorder = 2; GAD = 2; Mixed anxiety disorders = 1). Findings in this group were mixed. One study reported significantly lower CMT thickness in panic disorder patients as compared to controls (p = .02) (Baykara et al., 2022), though this study was excluded from meta-analysis due to overlapping controls with a more recent publication by the same group. The more recent study observed lower macular volumes in GAD patients (p = .016) (Baykara et al., 2024), while lower quadrant-specific macular thickness outside the CMT was reported in unmedicated GAD cases (p < .05)(Ozisik and Kiraz, 2023). In contrast, Acan et al. (2024) reported no significant differences between controls and anxiety disorder cases, consisting of individuals diagnosed with specific phobia, social anxiety disorder and agoraphobia, panic disorder and GAD (Acan et al., 2024). However, a more recent 2025 study showed that patients with panic disorder exhibited greater choroidal thickness alongside lower thickness in the RNFL, GCL and IPL relative to controls (Kalenderoglu et al., 2025). Notably, this study observed RNFL thickness increased in the same patients following 4 weeks of pharmacotherapy, suggesting these changes may be state-dependent. Overall, results suggest possible localised differences in the macula, but no consistent evidence of CMT differences in anxiety disorders.

#### Depression

Thirteen studies published between 2016 and 2025 investigated retinal differences in patients with major depressive disorder (MDD) with mixed findings. Several studies reported lower GCIPL (Ganglion Cell – Inner Plexiform Layer) thickness in cases relative to controls (ElShaarawi et al., 2022; Friedel et al., 2025), with small but significant correlations between depression severity and lower GCIPL and RNFL thickness (r = −0.20; p < .05) (Jung et al., 2020; Liu et al., 2022b). Repetitive transcranial magnetic stimulation treatment was associated with longitudinal RNFL thickening in treatment-resistant MDD (Genc et al., 2019), while SSRI use was associated with greater GCIPL values, but not with differences in the RNFL (Jung et al., 2020). Other structural differences included lower thickness in the Outer Nuclear Layer, which negatively correlated with depressive symptom severity(Friedel et al., 2025), reduced thickness in specific GCIPL quadrants (Liu et al., 2021a; Liu et al., 2021b; Wang et al., 2024) and lower choroidal thickness measures (Turhan et al., 2024). However, multiple studies reported no significant OCT differences in patients with depression (Liu et al., 2022a; Sönmez et al., 2017; Xiao et al., 2024; Yildiz et al., 2016). Overall, lower GCIPL thickness was the most consistent finding, with some evidence linking reduced retinal thickness to disease severity, though findings remain heterogeneous.

#### SUD

Fifteen studies published between 2019 and 2024 investigated retinal differences across different substances (Alcohol = 3; Cannabis = 2; Cocaine = 1; Methamphetamine = 3; Opioid = 5; Multiple Substance Use [MSUD] = 1). Alcohol use disorder, a known neurotoxin(Stuart et al., 2023), was associated with lower thickness across multiple RNFL quadrants (Liu et al., 2021a), as well as reduced macular thickness and lower GCL thickness (Álvarez-Sesmero et al., 2019). One study excluded from meta-analysis due to an overlapping control group also reported lower RNFL and GCL thickness in individuals diagnosed with alcohol use disorder (Orum and Kalenderoglu, 2020). Cannabis findings were inconsistent: one study reported lower RNFL thickness (Dayi et al., 2020), while another reported greater RNFL thickness, suggested by the investigators as possibly reflecting the neuroprotective effects of cannabis (Kalenderoglu et al., 2020). This discrepancy may stem from the differences in participant abstinence, as Kalenderoglu et al. (2020) assessed patients in prolonged remission (mean ∼ 121 days), whereas Dayi et al. (2020) evaluated individuals without a defined period of abstinence. Cocaine users showed lower RNFL thickness across several quadrants (Gemelli et al., 2019).

Methamphetamine studies showed variable results, likely influenced by the timing of assessment relative to active use. Kaya and Kaya (2023) and Demir et al. (2022) both assessed participants on the first day of hospitalisation, capturing the acute phase of withdrawal. However, results were inconsistent: Kaya and Kaya (2023) observed greater thickness in multiple RNFL quadrants consistent with acute inflammatory oedema (Kaya and Kaya, 2023), while Demir et al. (2022) observed no differences between cases and controls, which may reflect an intermediate phase where acute thickening masks chronic neurodegeneration. In contrast, Talebnejad et al. (2020) reported quadrant-specific lower thickness with dose-dependent correlations, though they did not specify the duration of abstinence for their cohort.

Opioid studies were also highly heterogeneous: some found no changes (Karadere et al., 2020), others reported lower RNFL, GCL and CMT thickness (Khalil et al., 2023; Özsoy et al., 2023), while studies from the same group reported transient RNFL quadrant thickening before treatment with buprenorphine/naloxone, which decreased post treatment (Orum and Kalenderoglu, 2021; Orum et al., 2022). Specifically, Orum et al. (2022) observed this greater thickness in subjects who were confirmed urine-positive at the time of imaging, indicating that acute intoxication may produce transient retinal changes distinct from lower thickness seen in prolonged remission.

One study investigated retinal differences in patients with MSUD. In contrast to the lower thickness observed in alcohol use, patients with MSUD showed greater thickness in the global RNFL alongside lower thickness in the CMT. Notably, negative correlations were found between RNFL thickness and addiction severity (Kaya and Kaya, 2024).

### Meta-Analysis Results

#### Overall meta-analysis

This primary analysis pooled 145 effect sizes from 25 unique studies, aggregating all available retinal thickness measures across all assessed layers and quadrants examining retinal thickness measures in patients with anxiety, depression, or SUD, compared to healthy controls. No significant differences in retinal thicknesses were observed between cases and controls (pooled standardised mean difference (SMD) = −0.20; 95% CI [−0.53, 0.14]; p = .244) (Table 2). However, there was significant heterogeneity across the studies (Q = 756.57, p < .001, tau^2^ = 0.69). Please refer to the Supplementary Figure S1 for additional model results.

**Table 2:**
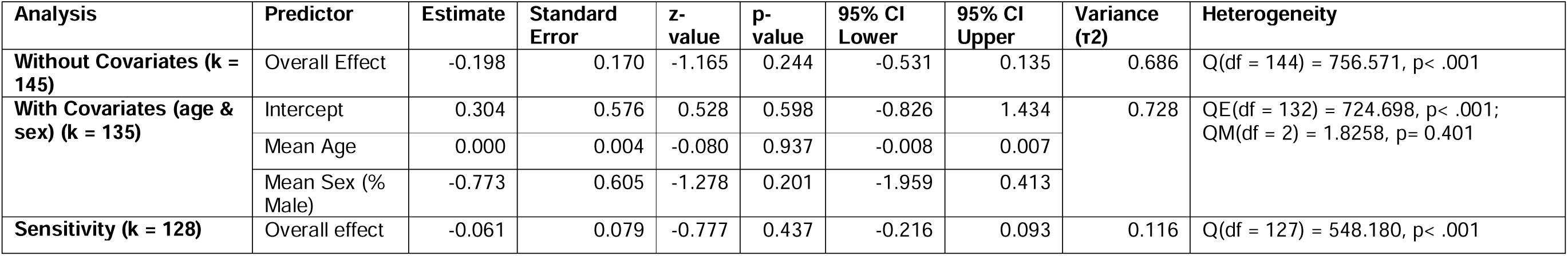
Overall Meta-Analysis: Combined Results for All Cases vs. All Controls.

| Analysis | Predictor | Estimate | Standard Error | z-value | p-value | 95% CI Lower | 95% CI Upper | Variance (τ<sup>2</sup>) | Heterogeneity |
| --- | --- | --- | --- | --- | --- | --- | --- | --- | --- |
| **Without Covariates (k = 145)** | Overall Effect | -0.198 | 0.170 | -1.165 | 0.244 | -0.531 | 0.135 | 0.686 | Q(df = 144) = 756.571, p< .001 |
| **With Covariates (age & sex) (k = 135)** | Intercept | 0.304 | 0.576 | 0.528 | 0.598 | -0.826 | 1.434 | 0.728 | QE(df = 132) = 724.698, p< .001;<br>QM(df = 2) = 1.8258, p= 0.401 |
|  | Mean Age | 0.000 | 0.004 | -0.080 | 0.937 | -0.008 | 0.007 |  |  |
|  | Mean Sex (% Male) | -0.773 | 0.605 | -1.278 | 0.201 | -1.959 | 0.413 |  |  |
| **Sensitivity (k = 128)** | Overall effect | -0.061 | 0.079 | -0.777 | 0.437 | -0.216 | 0.093 | 0.116 | Q(df = 127) = 548.180, p< .001 |

#### Sensitivity Analysis

To test the robustness of the primary findings, a sensitivity analysis was conducted. First, a manual leave-one-out cross-validation was conducted, excluding one study at a time to determine if any single study disproportionately influenced the pooled estimate. The analysis demonstrated high stability: the pooled estimate fluctuated minimally (−0.24 to −0.05) and no single study exclusion altered the non-significant finding (p > .05).

Secondly, forest plots were visually inspected to identify effect sizes that were extreme outliers. Two such outliers were identified: Talebnejad et al. (2020), which reported an extreme effect size (SMD = −4.50) associated with lower thickness in chronic methamphetamine use, and Orum et al. (2022), which reported longitudinal retinal thickening with acute opioid intoxication. This pattern distinctly contrasted from the lower thickness trends observed in the broader cohort, as evidenced across the global RNFL (SMD range +0.18 to +0.32) up to SMD +0.99 in the superior quadrant, which normalised post-treatment. Excluding Orum et al. (2022) alone (k = 129), Talebnejad et al. (2020) alone (k = 144), or both studies simultaneously (k = 128; SMD = −0.06; 95% CI[−0.22, 0.09]; p = .437) did not alter the non-significant results, confirming that the primary findings were not driven by these outlying studies.

#### Moderator Analyses

Given the significant heterogeneity identified in the primary analysis, a series of exploratory meta-regressions were performed across all eligible studies to investigate sources of heterogeneity, specifically the moderating effects of age and sex and testing for an age × diagnosis interaction. The included studies with age data (n = 24) consisted of young to middle-aged adults, with an overall weighted mean age of 34.93 years (SD = 12.35; range 23.08 - 62.50 years). The diagnostic subgroups were well matched, with similar mean ages for anxiety (38.07 years; SD = 10.19), depression (37.11 years; SD = 13.52) and SUD (34.33 years; SD = 14.16).

Using a stepwise approach, the primary model controlling for age and sex (Table 2) found no association between age (p = .937) or sex (p = .201) and retinal thickness across the sample. To test for an age × diagnosis interaction, comparative meta-regressions (Table 4) found no relationship between age and retinal thickness by diagnosis, relative to healthy controls (QM(df = 8) = 6.01, p = .647). The age slopes for depression (SMD = 0.15, p = .826) and SUD (SMD = 0.09, p = .894) were not significantly different from the anxiety subgroup. Similarly, no significant sex × diagnosis interactions were observed. Therefore, our analyses found no evidence that the association between psychiatric diagnosis and retinal thickness is moderated by age or sex.

### Subgroup - Specific Analyses

#### Anxiety Disorders

Three studies were included for meta-analysis (Figure 2), with Central Macular Thickness (CMT) being consistently reported across all studies. Pooled estimates showed no significant differences between patients with anxiety disorders and healthy controls (pooled SMD = −0.06; 95% CI[−0.35, 0.22]; p = .671), with minimal heterogeneity across studies (Q(df = 2) = 0.76, p = .683) (Table 3). This subgroup showed no evidence of publication bias, based on visual inspection of the funnel plot and non-significant Egger’s test (p = .615). However, this is underpowered due to the low number of included studies (k =3).

**Figure 2:**
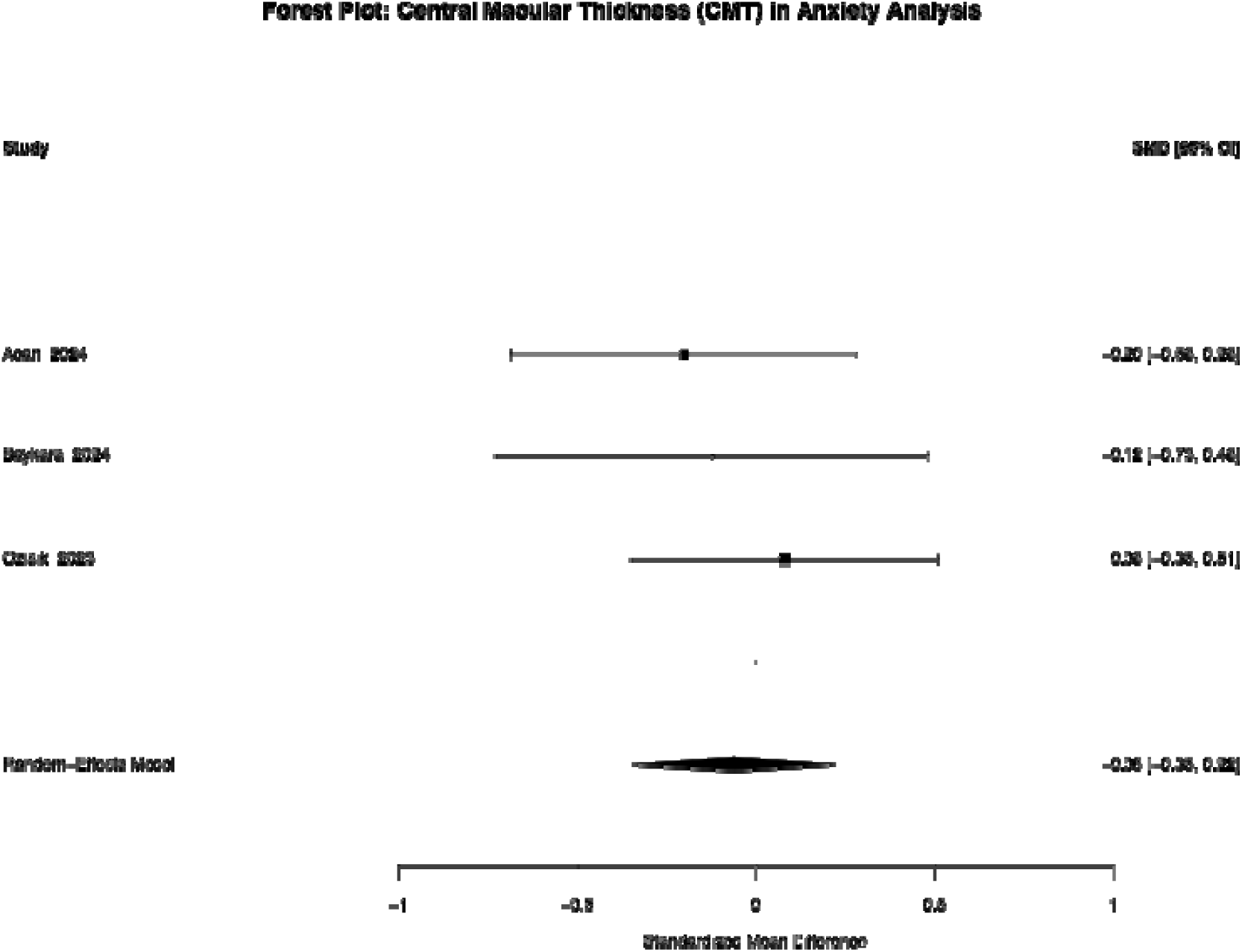
Anxiety Meta-Analysis Forest Plot

**Table 3:**
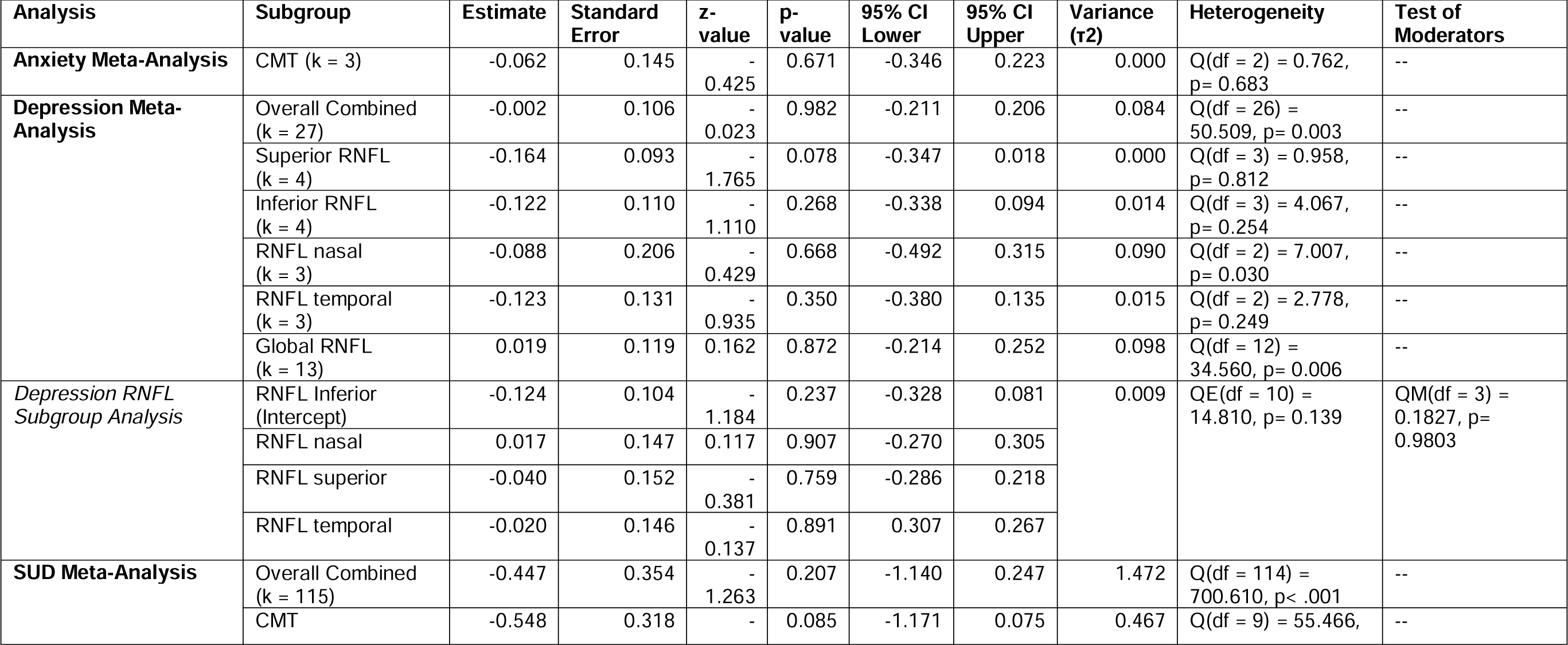
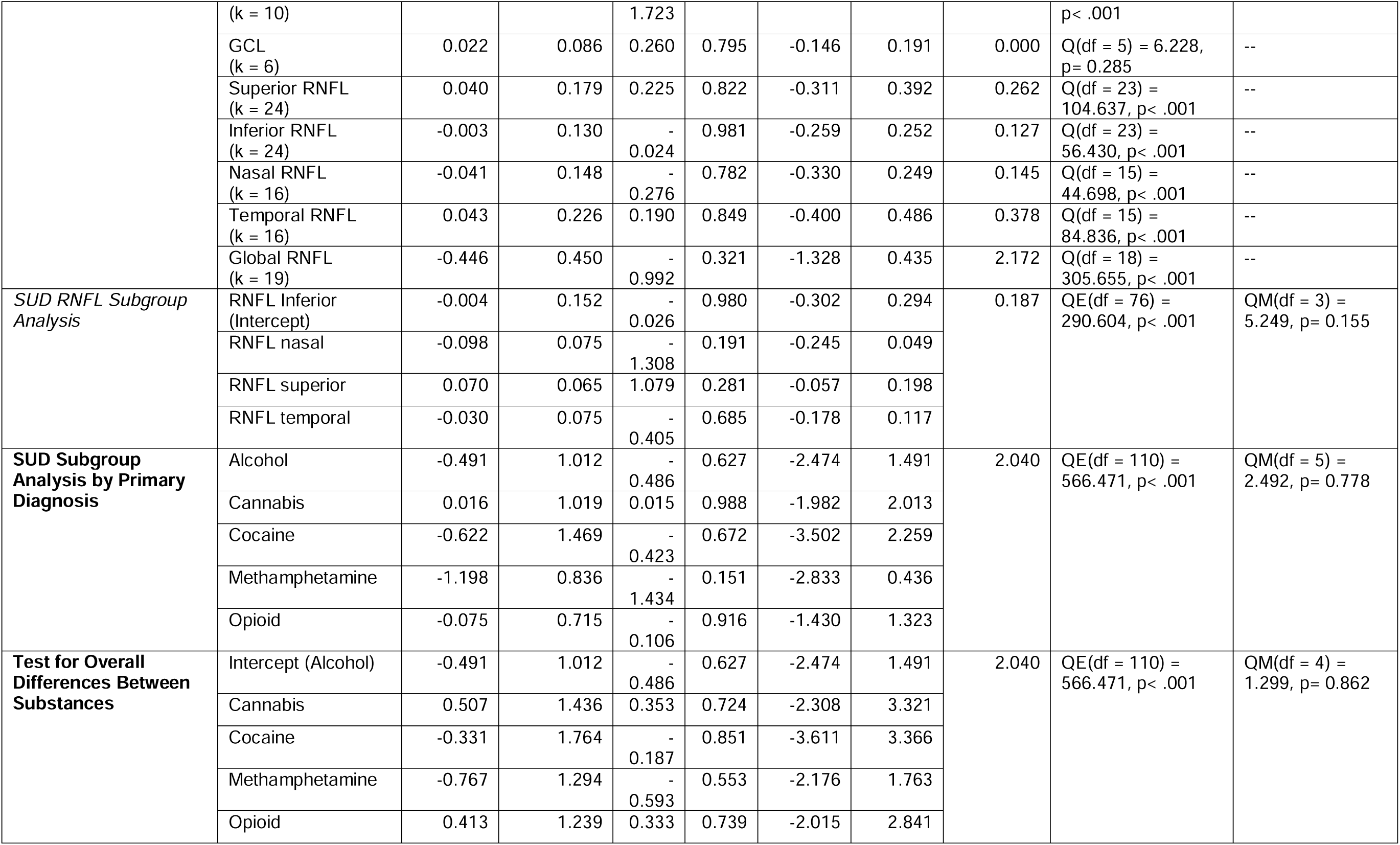
Subgroup Analyses.

**Table 4:**
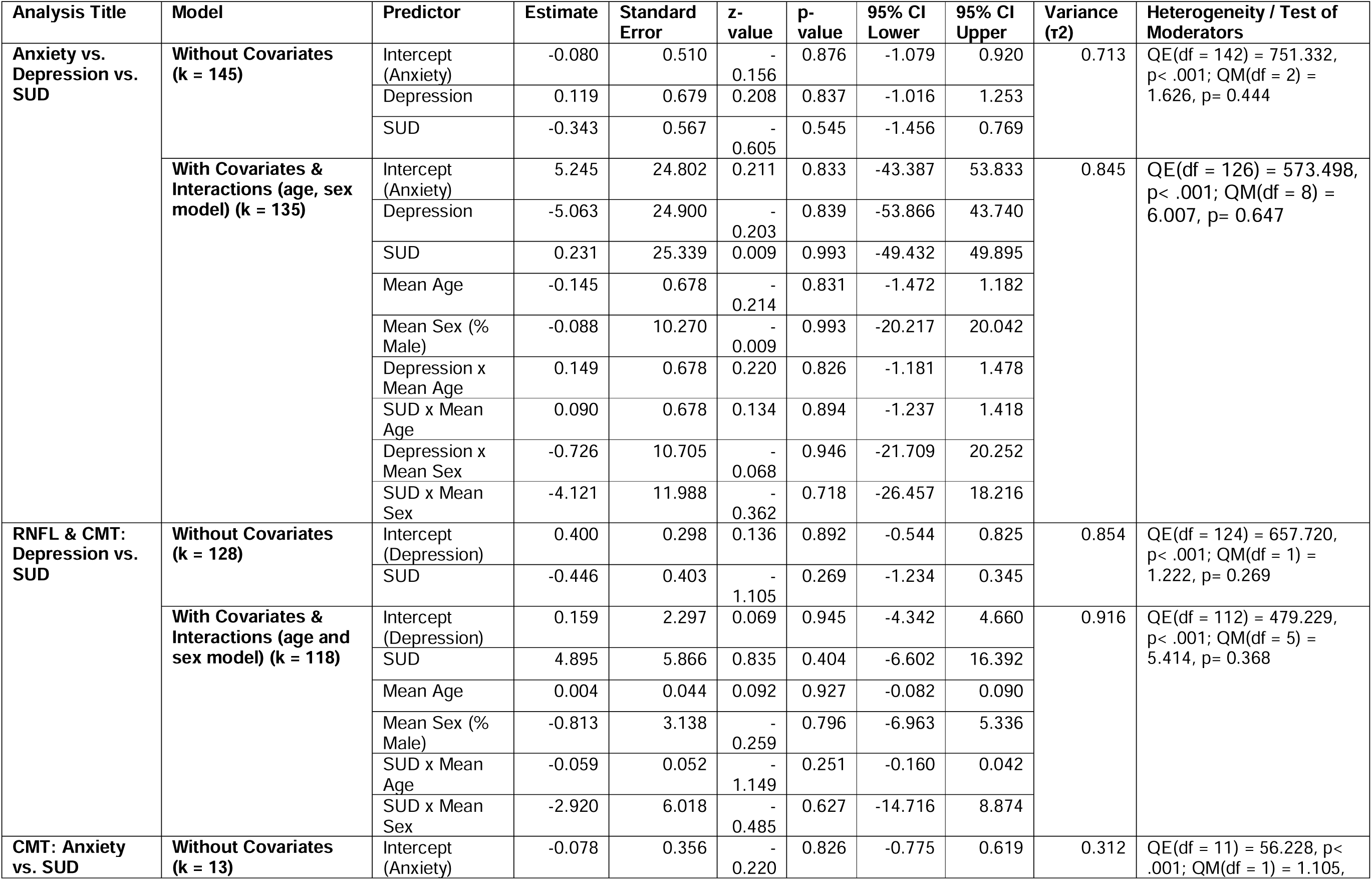
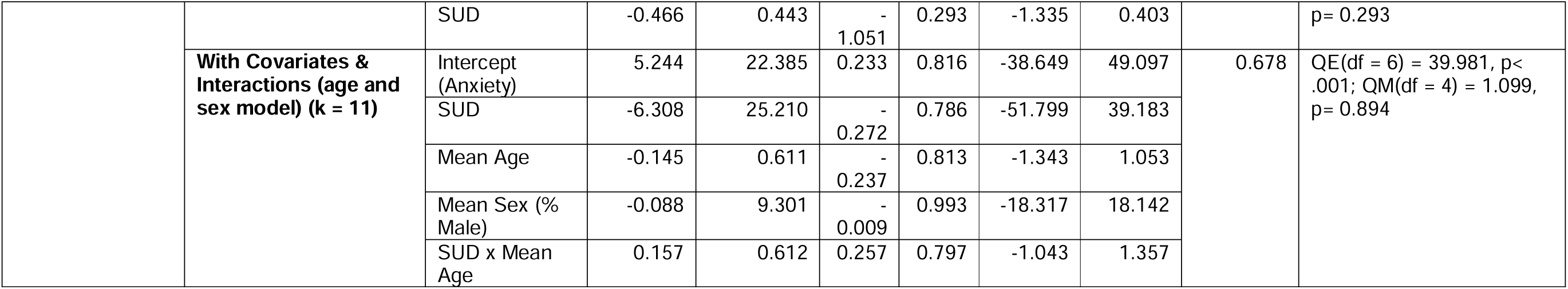
Comparative Meta-Analysis Results.

#### Depression Subgroup

A total of 27 effect sizes from 10 unique studies were included for the depression subgroup, with both overall retinal thickness and individual RNFL quadrants analysed (Figure 3). The pooled overall estimate failed to detect evidence for group differences (pooled SMD 0.00; 95% CI[−0.21, 0.21]; p= .982) between depression cases and controls, though heterogeneity was significant (Q(df = 26) = 50.51; p = .003) (Table 3).

**Figure 3:**
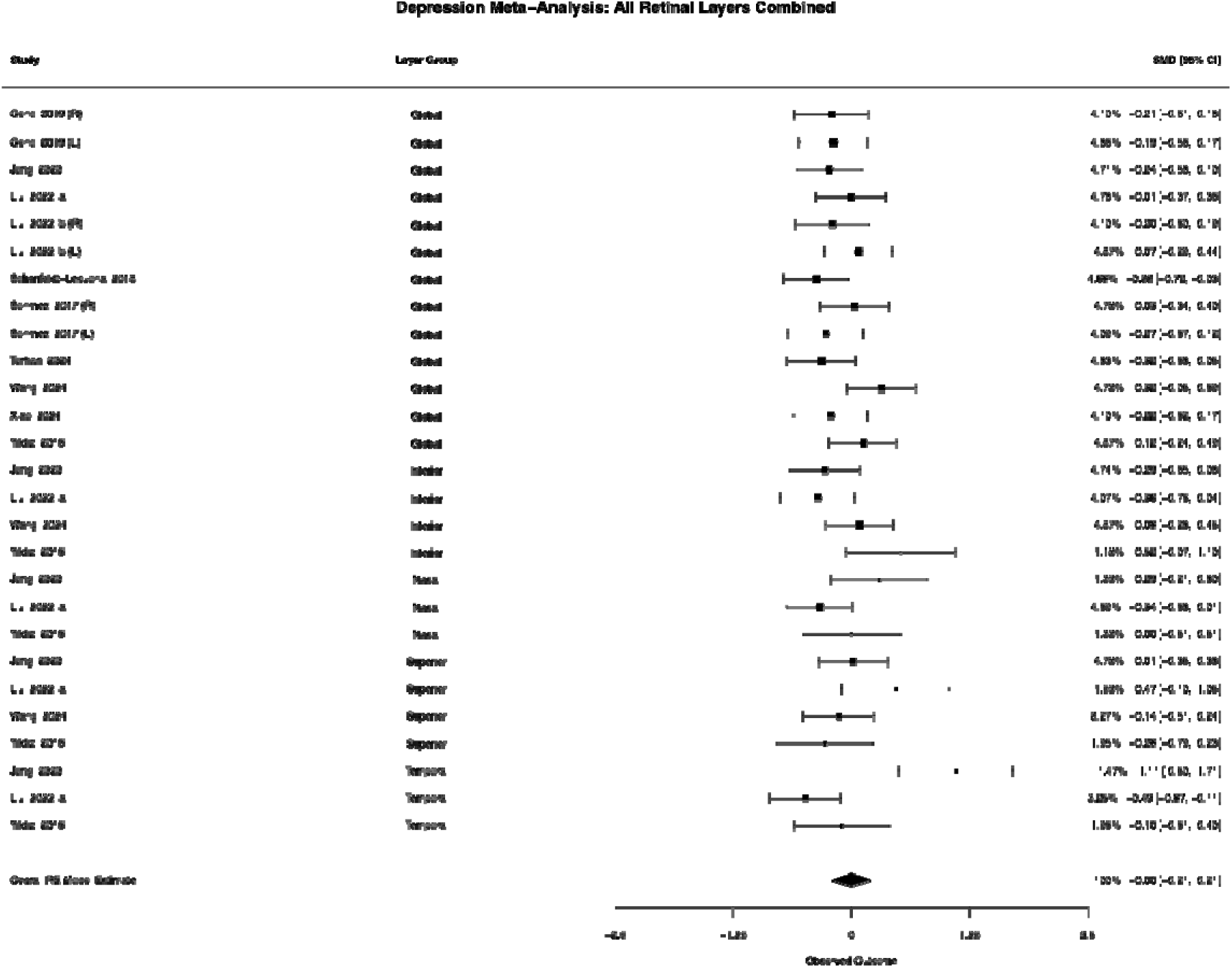
Depression Meta-Analysis Forest Plot

Quadrant-specific RNFL analyses, based on data availability, were also non-significant for the inferior (pooled SMD = −0.12; p = .268), nasal (pooled SMD −0.09; p = .668), and temporal quadrants (pooled SMD = −0.12; p = .350) as well as the global RNFL (pooled SMD = 0.02; p = .872) (Table 3). The superior quadrant showed a trend towards lower thickness, although this did not reach significance (pooled SMD = −0.16; p = .078). Quadrant comparison analysis found that lower RNFL thickness associations are not localised to a specific quadrant in depression (QM(df = 3) = 0.18; p = .980).

Analysis of both the device-measured global RNFL and the calculated composite of all RNFL quadrants (overall RNFL) revealed evidence of small study effects. This was demonstrated by a significant standard error (sei) coefficient (p < .05), showing that studies with lower precision reported more extreme effect sizes as compared to larger, more precise studies. This asymmetry suggests potential publication bias influence. In contrast, individual quadrant analyses showed no clear evidence of bias, although these models were limited by a small number of studies.

#### Substance Use Disorder Subgroup

The largest subgroup analysis included 115 effect sizes from 12 studies for SUD. Overall, retinal thickness measures did not differ significantly between individuals with SUD and healthy controls (pooled SMD = −0.45; 95% CI[−1.14, 0.25]; p = .207), though heterogeneity was significant (Q(df = 114) = 700.61; p < .001; tau^2^ 1.47) (Table 3). The summary estimates by substances are shown in Figure 4. Full forest plots for each retinal layer are provided in the Supplementary Figure S3.

**Figure 4:**
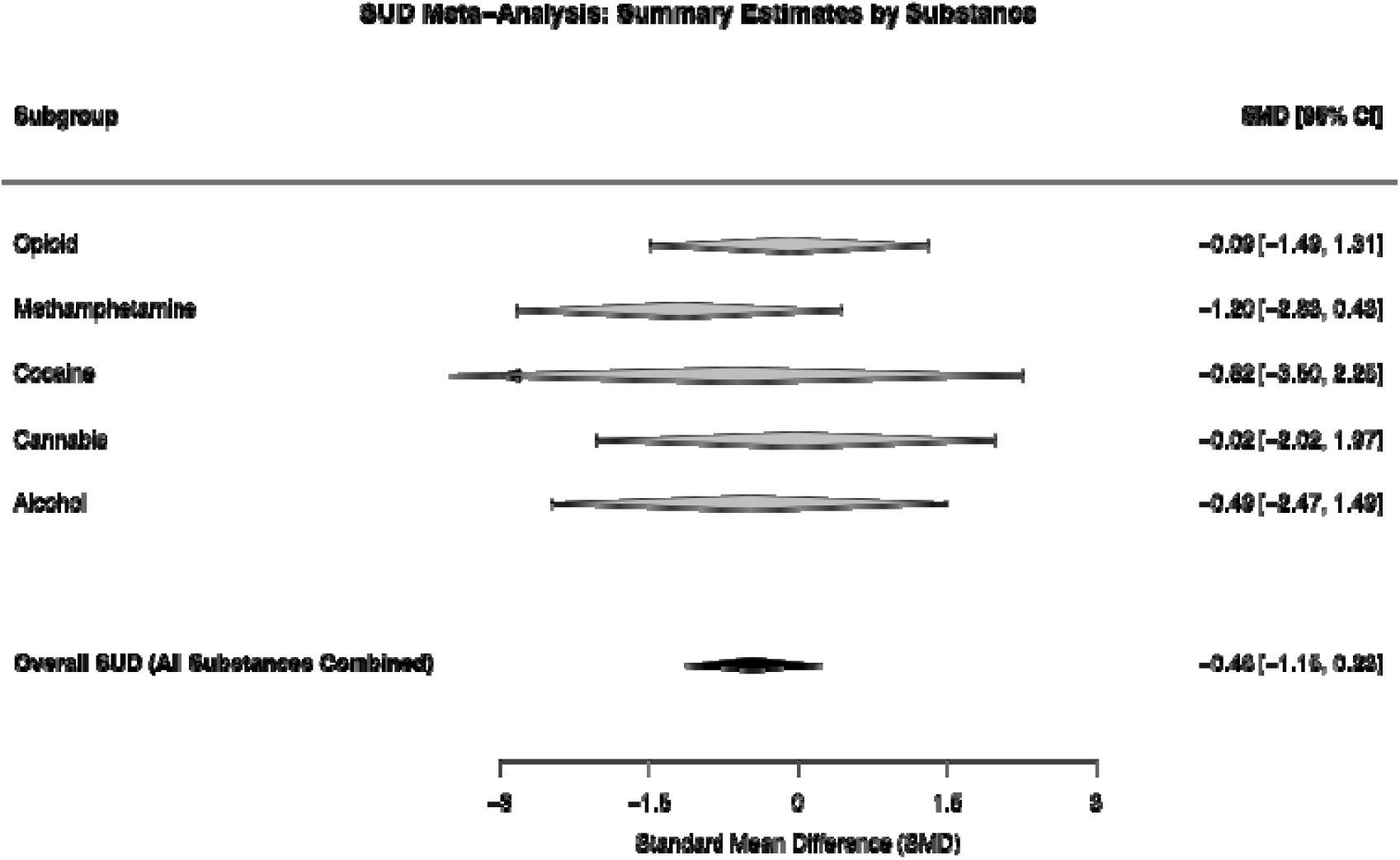
SUD Meta-Analysis Summary Forest Plot of Retinal Thickness. Pooled standardised mean differences (SMD) of retinal thickness measures are shown for each substance group in Figure 4. The overall pooled estimate indicates no significant differences between cases and controls. Detailed forest plots are available in Supplementary Figures S3.

Within SUD, layer-specific analysis showed a trend toward lower CMT thickness that failed to reach statistical significance (pooled SMD −0.55; 95% [CI −1.17, 0.08], p = .085). No significant effects were detected for the GCL (estimate = 0.02; p = .795), any RNFL quadrants (all p > .78) or global RNFL (estimate = −0.45; p = .321) (Table 3). Similar to the depression findings, a formal comparison between the RNFL quadrants (superior, inferior, nasal and temporal) found no significant differences (QM(df=3) = 5.25, p = .155), suggesting no specific quadrant is preferentially affected in SUD.

When stratified by substance type, no significant differences were found for alcohol, cannabis, cocaine, methamphetamine or opioid use when compared to controls. Subsequent analysis confirmed that substance type did not significantly moderate these results (QM(df = 5) = 2.49, p = .778), suggesting the effect is consistent across diagnoses (Table 3). These results should be interpreted with caution due to the extremely small number of studies contributing to each subgroup. Significant publication bias was present for the overall SUD analysis (p = .002) and CMT analysis (p = .021), indicating an association between the precision of the effect size reported by a study (a function of study sample size) and the size of the effect reported.

#### Publication Bias

Publication bias was assessed using Egger’s regression test for funnel plot asymmetry based on the standard error (sei) coefficient. Significant asymmetry was detected in the primary overall analysis (k = 145, p = .004) (Figure 5, A). However, in the analysis excluding the two visually identified outliers (k = 128), the test for funnel plot asymmetry was no longer significant (p = .241) (Figure 5, B). This suggests that the observed asymmetry may be influenced by these two distinct outliers (Orum et al., 2022; Talebnejad et al., 2020) representing clinical extremes, although the presence of broader small-study effects cannot be fully excluded.

**Figure 5:**
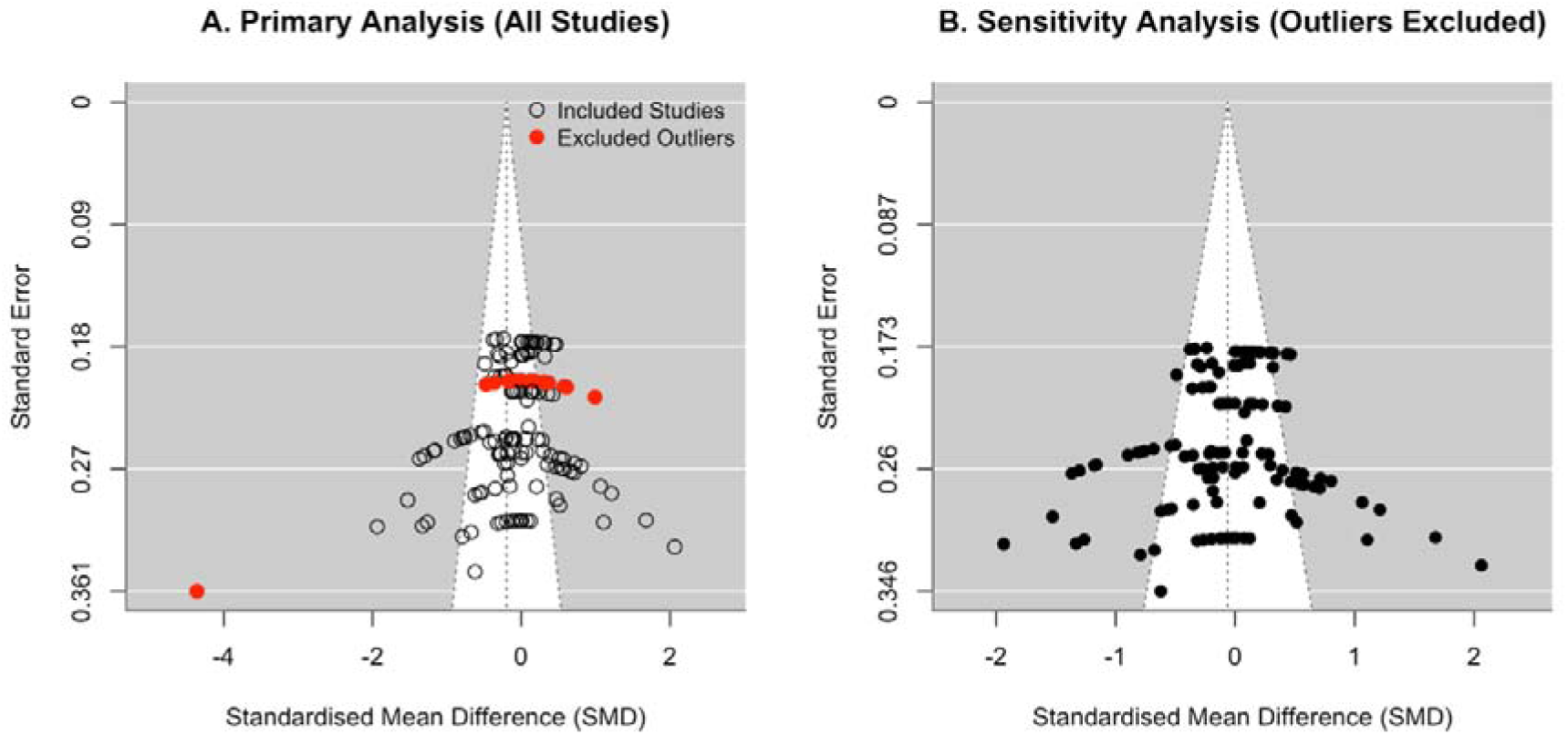
Funnel Plots for Publication Bias. (A) Primary analysis showing asymmetry driven by outliers (red dots). (B) Sensitivity analysis after removing outliers (Orum et al., 2022; Talebnejad et al., 2020), showing restored symmetry.

## Discussion

This systematic review and meta-analysis evaluated retinal structural differences across patients with anxiety, depression, and substance use disorders (SUD). Although the majority of individual studies reported differences between cases and controls in at least one outcome (4 of 5 in the anxiety group, 9 of 13 in the depression group, and 13 of 15 in the SUD group), the pooled meta-analysis did not identify a consistent overall effect. This stands in contrast to the robust association observed in neurodegenerative research, where meta-analyses have consistently established lower retinal thickness as a reliable predictor of cognitive decline and incident dementia (Chan et al., 2019; Coppola et al., 2015; Thomson et al., 2015), which has been further supported by large-scale prospective cohort studies (Ko et al., 2018; Mutlu et al., 2018). These findings suggest that while retinal imaging remains a theoretically compelling target for psychiatric screening, the current evidence, characterised by high heterogeneity, methodological variability, and potential publication bias, is insufficient to support its clinical application in these psychiatric conditions.

Our primary aim was to determine if retinal abnormalities are present in anxiety, depression, and SUD. No single retinal layer had sufficient, comparable data to be included in the meta-analysis across all disorders. In patients with major depression, smaller GCIPL thickness estimates were consistently reported in all 4 of the studies that reported on this layer (ElShaarawi et al., 2022; Friedel et al., 2025; Jung et al., 2020; Turhan et al., 2024). Unfortunately, due to data reporting inconsistencies in the primary studies, these effects could not be pooled in our meta-analysis. RNFL results were mixed, with only 3 out of the 13 papers reporting significantly lower thickness estimates (Liu et al., 2022b; Liu et al., 2021b; Wang et al., 2024). This consistent finding with respect to the GCIPL aligns with the hypothesis that the macular region, which has a high density of retinal ganglion cells and high metabolic demand, may be particularly vulnerable to neurodegenerative disease and inflammatory processes (Baltan et al., 2010; Celik et al., 2016; Cheung et al., 2021; Ruggeri et al., 2025).

In contrast, findings in anxiety disorders were scarce and heterogeneous, with no reproducible pattern. However, individual studies offered limited evidence that these measures may be state-dependent. For example, one study observed longitudinal RNFL thickening following pharmacotherapy, findings that, like those in the SUD literature, suggest that treatment status and acute symptoms management may influence retinal morphology (Kalenderoglu et al., 2025).

A significant conceptual challenge in interpreting findings across all three disorders is the cross-sectional nature of the current literature. Without longitudinal studies tracing the natural course of depression and anxiety in relation to retinal health, it remains unclear whether observed retinal differences represent pre-existing psychiatric vulnerability, or a consequence of the illness itself. In the context of SUD, this challenge is further compounded by the potential for direct neurotoxicity from exogenous substances. Unlike anxiety and depression, where retinal differences are hypothesised to reflect endogenous neuroinflammatory and neurodegenerative processes, substances such as alcohol (Stuart et al., 2023) and methamphetamine (Jayanthi et al., 2021) are known neurotoxins that may exert direct chemical insults on the retina. Therefore, it remains unclear whether the observed lowered thickness represents a pre-existing psychiatric vulnerability or a consequence of chronic substance exposure. To date, no prospective studies currently track these measures longitudinally across any of these patient groups. Nevertheless, the ability of OCT to quantify neurodegeneration in this patient population, after consideration of these factors, as well as variables such as duration of abstinence, remains clinically relevant for monitoring the systemic impact of long-term substance use.

Findings from substance use disorder studies were also highly divergent across the relatively large number of effect sizes reported for this group. Within this context, while alcohol (n = 3) and cocaine (n = 1) use were generally associated with lower thickness, cannabis and opioid use produced mixed results, with some reported lower thickness (cannabis n = 1; opioid n = 2) and others greater thickness (cannabis n = 1; opioid n = 1) in the RNFL, with an opioid study also showing pre-and post-treatment changes(Orum et al., 2022). Methamphetamine studies also showed mixed results in the narrative review, reporting both greater and lower thickness across individual studies.

There is evidence suggesting that some of the heterogeneity in outcomes reported for studies of OCT in substance users may be driven by the phase of substance use, rather than systemic bias. For instance, Orum et al. (2022) recruited participants who were urine positive for opioids at the time of scanning, showing longitudinal retinal thickening that subsequently normalised post-treatment, likely suggesting acute inflammation masking chronic neurodegeneration. This masking effect may also explain why Demir et al. (2022) observed no significant differences despite assessing patients on the first day of hospitalisation. Conversely, Talebnejad et al. (2020) captured chronic methamphetamine users, reporting a dose-dependent correlation with reduced retinal thickness. The potential influence of chronicity on diagnosis is further underscored by findings in Multiple Substance Use Disorder, where greater RNFL thickness is reported alongside lower macular thickness, supporting the more general observation that retinal structures are dynamic, with their presentation likely to differ based on substance type, recency of use and addiction severity.

Notably, the inflammatory mechanism may not apply to all substances. Kalenderoglu et al. (2020) suggested that the greater thickness observed in their cohort of cannabis users may reflect a neuroprotective effect. This stands in contrast to the thickening observed in opioid and methamphetamine users, which appears more likely to be driven by an acute neuroinflammatory response or oedema, rather than neuroprotection (Jayanthi et al., 2021)

Our second aim was to evaluate consistency across the three disorders. When comparing the three disorders directly, no significant differences in effect sizes were found. As noted above, no single layer was consistently affected across all three conditions, however, given the scarcity and heterogeneity of the available data, the current literature may simply be underpowered to detect a shared pathophysiological biomarker.

We also explored whether associations of retinal thickness with chronological age varied by disorder, as tested by an age × diagnosis moderator analysis. Our analysis did not find an association between age and retinal thickness. The stepwise investigation confirmed this, ultimately finding no robust evidence for an age × diagnosis interaction across any of the models. This finding, as well as the lack of evidence in this study for a main effect of age on retinal thickness however, may reflect the sample’s young demographic profile, with the overall sample weighted mean age of 34.93 (range 23.08 - 62.50 years). Previous studies have provided robust evidence for an effect of age in the neurosensory retina (e.g. RNFL, GCIPL) as part of the normal ageing process (Harwerth et al., 2008; Leung et al., 2012; Mwanza et al., 2011; Sung et al., 2009). In such a young population with a limited age range, this effect may be statistically undetectable and possibly masked by the heterogeneity observed in the studies.

Beyond chronological age, the duration of illness remains a critical factor that likely influences the progression of retinal structural changes. A longer duration of illness represents a greater cumulative burden of the disorder, which may be the primary driver of accelerated retinal thinning in the context of exposure to neuroinflammation. For instance, while Yildiz et al. (2016) reported no significant overall differences between MDD cases and healthy controls, the duration of the depressive episode was negatively correlated with GCIPL and nasal RNFL thickness. However, as noted in our review, few studies consistently reported or controlled for illness duration.

### Study Limitations and Strengths

The strengths of this study include the adherence to the PRISMA guidelines and the use of a robust multilevel statistical model to address the non-independence of data arising from multiple measurements from the same study participants. This method prevents the artificial inflation of statistical significance that can occur when dependent effect sizes are treated as independent, ensuring more reliable variance estimation(Gooty et al., 2019). Furthermore, the removal of two identified outliers resolved the statistical evidence of publication bias.

However, the interpretation of findings is limited by the considerable heterogeneity in the source studies, rather than the review process itself. Methodologically, interpretation is difficult due to the diversity of instrumentation and acquisition methods. Multiple different OCT machines were used in the various studies (e.g. Heidelberg Spectralis vs. Zeiss Cirrus), which utilise different segmentation algorithms and sector definitions. Our analysis had to combine measurements from various retinal sub-sectors to maximise data inclusion. While necessary, this manual aggregation likely introduced additional variance and contributed to observed heterogeneity. Furthermore, inconsistencies in reporting were common in the source studies. Few reported controlling for confounders such as intraocular pressures (Nishida et al., 2022), axial length (Nasir et al., 2025) and optic disc size, all of which can inherently affect thickness measures. Clinically, psychiatric diagnoses used a range of instruments, while illness duration, severity and treatment status were inconsistently described.

Beyond these inconsistencies, several factors limit the generalisability and statistical power of these findings. Regional clustering of studies was also evident, with a disproportionate number of studies conducted in Turkey and China. Several studies were limited by small sample sizes, increasing susceptibility to chance or restricted their samples of one sex. The young demographic profile may have made an age × diagnosis interaction statistically undetectable, as the effects of normal ageing are less pronounced in younger populations. Finally, while the number of eligible studies included for meta-analysis is substantial, the limited number of studies identified within specific subgroups remains a constraint, which limits the statistical power of the study and the ability to explore sources of heterogeneity through moderator analyses.

Moreover, the interpretation of the SUD literature is further complicated by the high number of reported measures relative to the number of studies included in the meta-analysis (115 effect sizes across 12 papers). This pattern may increase the risk of false positives at the individual study level, potentially explaining why specific papers reported ‘significant’ differences that were not replicated in the present pooled analysis.

Consequently, while the pooled meta-analysis revealed no significant differences, the results should be interpreted with caution, due to the observed heterogeneity, which is arguably the most important finding of this study.

### Implications for further research

Based on this comprehensive meta-analysis, OCT-derived thickness measures are not currently supported by evidence as reliable biomarkers for diagnosing or monitoring anxiety, depression, or substance use disorders. The heterogeneous literature in this area is one of the major contributing factors. However, the success of retinal imaging in Multiple Sclerosis (Petzold et al., 2021) and Alzheimer’s disease (Cheung et al., 2021) suggests that the potential for objective psychiatric biomarkers remains high.

The theoretical rationale for retinal imaging in psychiatry remains intact, through the shared embryological and anatomical ties between the retina and central nervous system (Foster and Khaw, 2009; London et al., 2013).However, significant improvement in study methodologies in the three psychiatric disorders of interest is required to realise this potential. Future research should follow the neurological blueprint to include large-scale, multi-centred, prospective studies with standardised protocols for OCT imaging and diagnostic criteria. Minimum reporting standards should be established and mandated, covering medication status, illness duration, axial length, intraocular pressure, optic disc area and OCT signal strength, modelled on existing neuroimaging reporting frameworks (Youm et al., 2012). Standardised and consistent reporting of specific retinal layers is also critical to facilitate future meta-analyses. For example, the consistent narrative finding of lower GCIPL thickness in depression observed in this review could not be meta-analysed due to reporting variation. Prospective designs would help to disentangle whether retinal changes are markers of active illness, or a consequence of chronic disease or treatment exposure, while standardised and consistent reporting of retinal layers across studies would facilitate meta-analyses.

In substance use disorder research specifically, study designs must account for retinotoxicity and distinguish between the acute neuroinflammatory and chronic neurodegenerative phases.

Integration of OCT findings with other biomarkers may be valuable and could clarify shared mechanisms between neuropsychiatric conditions and neurodegenerative processes(Banihashemi et al., 2025; Petzold et al., 2022). Furthermore, specific investigations should be focused on the realm of anxiety disorders, as this area in particular is currently under-researched. Specific emphasis should be placed on the GCIPL and macular protocols, which showed the most consistent signal across depression studies. These areas may be more sensitive to neurodegenerative changes than global RNFL measures, warranting systematic evaluation in future large-scale studies. Exploring specific thinning patterns between RNFL quadrants may also be valuable, as evidenced by some studies in the depression subgroup, as well as substance-specific patterns in the SUD group. Ultimately, resolving the current inconsistencies will require collaborative efforts which move beyond single-centred studies towards rigorous, reproducible projects. The recently established ENIGMA Retina Working Group, with its aim to establish normative models of retinal thickness to quantify deviations from healthy ageing, exemplifies this direction (Kallen et al., 2024).

In conclusion, while lower retinal thickness estimates have been reported on in individual studies of common mental disorders, our meta-analysis demonstrates that the collective evidence is not robust. This field of research is characterised by inconsistent findings and potential publication bias, highlighting the need for more methodologically rigorous and standardised research before the clinical utility of retinal biomarkers in psychiatry can be established.

## Data Availability

Data extracted from included studies are available upon reasonable request to the corresponding author.

## Acknowledgements

The authors wish to honour the memory of our co-author, Dan J. Stein, who passed away during the preparation of this manuscript on 5 December 2025. His vision and guidance were fundamental to the conceptualisation of this study. His contribution to scientific research is invaluable.

## Competing interests

No competing interests

## Author statement

**MG:** Conceptualisation, Methodology, Software, Formal analysis, Investigation, Data Curation, Writing - Original Draft, Writing - Review & Editing, Visualisation, Project administration, Funding acquisition. **SK:** Investigation, Data Curation; Writing - Review & Editing. **JI:** Conceptualisation; Methodology, Supervision, Writing - Review & Editing. **DS†:** Conceptualisation, Writing - Review & Editing. **RC, WH, ZS, PF, MH, ZZ:** Writing - Review & Editing.

## Funding statement

The work reported herein was made possible through funding by the South African Medical Research Council through its Division of Research Capacity Development under the SAMRC Institutional Clinician Researcher Development Programme with funding received from the National Department of Health.

## Declaration of generative AI and AI-assisted technologies in the manuscript preparation process

During the preparation of this work, the author(s) used Gemini 2.5 and 3.0 Pro to assist in the generation and optimisation of R code for the meta-analysis, as well as for proofreading of the manuscript. Following the use of these tools, the author(s) reviewed and edited the generated content and take full responsibility for the integrity and accuracy of the final published work.

## Notes

### Competing Interest Statement

The authors have declared no competing interest.

### Clinical Protocols

https://www.crd.york.ac.uk/PROSPERO/view/CRD42024559542

### Funding Statement

This work was supported by the South African Medical Research Council through its Division of Research Capacity Development under the SAMRC Institutional Clinician Researcher Development Programme, funded by the National Department of Health South Africa. The funders had no role in the study design; collection, analysis, or interpretation of data; writing of the report; or decision to submit the article for publication.

## References

Uncategorized References

Acan, D., Baykan, H., Karahan, E., 2024. Evaluation of retinal and choroidal vascular structures in patients with anxiety disorder. Eur. J. Ophthalmol., 6. 10.1177/11206721241228620.

Álvarez-Sesmero, S., Povedano-Montero, F.J., Arias-Horcajadas, F., Marín-Mayor, M., Navarrete-Chamorro, P., Raga-Martínez, I., Rubio, G., López-Muñoz, F., 2019. Retinal nerve fiber layer in patients with alcohol use disorder. Applied Sciences. 9, 5331.

Baltan, S., Inman, D.M., Danilov, C.A., Morrison, R.S., Calkins, D.J., Horner, P.J., 2010. Metabolic vulnerability disposes retinal ganglion cell axons to dysfunction in a model of glaucomatous degeneration. J Neurosci. 30, 5644–5652. 10.1523/jneurosci.5956-09.2010.

Banihashemi, M.A., Krance, S.H., Ji, P.X., Koo, M., Ottoy, J., Swartz, R.H., Kertes, P.J., Hudson, C., Goubran, M., Black, S.E., 2025. Brain and retina in Alzheimer’s disease: Pathological intersections and estimates from imaging. Alzheimers Dement. 21, e70884. 10.1002/alz.70884.

Baykara, S., Kazğan, A., Yıldırım, H., Tabara, M.F., Kaşıkcı, H., Danacı Keleş, D., 2024. Retinal changes in generalized anxiety disorder patients. Int J Psychiatry Med. 59, 270–286. 10.1177/00912174231209771.

Baykara, S., Yıldırım, H., Kazğan, A., Tabara, M.F., Keleş, D.D., Gürok, M.G., Atmaca, M., 2022. Retinal Changes in Panic Disorder Patients. Psychiatry Res Neuroimaging. 324, 111496. 10.1016/j.pscychresns.2022.111496.

Bonnel, S., Mohand-Said, S., Sahel, J.-A., 2003. The aging of the retina. Experimental gerontology. 38, 825–831.

Bora, E., Fornito, A., Pantelis, C., Yücel, M., 2012. Gray matter abnormalities in Major Depressive Disorder: A meta-analysis of voxel based morphometry studies. Journal of Affective Disorders. 138, 9–18. 10.1016/j.jad.2011.03.049.

Celik, M., Kalenderoglu, A., Sevgi Karadag, A., Bekir Egilmez, O., Han-Almis, B., Şimşek, A., 2016. Decreases in ganglion cell layer and inner plexiform layer volumes correlate better with disease severity in schizophrenia patients than retinal nerve fiber layer thickness: Findings from spectral optic coherence tomography. Eur Psychiatry. 32, 9–15. 10.1016/j.eurpsy.2015.10.006.

Chan, V.T.T., Sun, Z., Tang, S., Chen, L.J., Wong, A., Tham, C.C., Wong, T.Y., Chen, C., Ikram, M.K., Whitson, H.E., Lad, E.M., Mok, V.C.T., Cheung, C.Y., 2019. Spectral-Domain OCT Measurements in Alzheimer’s Disease: A Systematic Review and Meta-analysis. Ophthalmology. 126, 497–510. 10.1016/j.ophtha.2018.08.009.

Cheung, C.Y., Mok, V., Foster, P.J., Trucco, E., Chen, C., Wong, T.Y., 2021. Retinal imaging in Alzheimer’s disease. J Neurol Neurosurg Psychiatry. 92, 983–994. 10.1136/jnnp-2020-325347.

Coppola, G., Di Renzo, A., Ziccardi, L., Martelli, F., Fadda, A., Manni, G., Barboni, P., Pierelli, F., Sadun, A.A., Parisi, V., 2015. Optical coherence tomography in Alzheimer’s disease: a meta-analysis. PloS one. 10, e0134750.

Dayi, A., Dayi, Ö., Kurt, E., Yorguner, N., 2020. [Optical Coherence Tomography Findings in Cannabis Users]. Turk Psikiyatri Derg. 31, 239–243. 10.5080/u25205.

Demir, B., Ozsoy, F., Kepenek, I., Altindag, A., 2022. Examination of optical coherence tomography findings in patients with methamphetamine use disorder. J. Addict. Dis. 40, 278–284. 10.1080/10550887.2021.1983294.

Demirkaya, N., Van Dijk, H.W., Van Schuppen, S.M., Abràmoff, M.D., Garvin, M.K., Sonka, M., Schlingemann, R.O., Verbraak, F.D., 2013. Effect of age on individual retinal layer thickness in normal eyes as measured with spectral-domain optical coherence tomography. Investigative ophthalmology & visual science. 54, 4934–4940.

ElShaarawi, M.S., Gaafar, A.A., Shalaby, H.S., Abdelgawad, R.H.A., 2022. Optical coherence tomography changes in Egyptian patients with major depressive disorder. J. Egypt. Ophthalmol. Soc. 115, 169–174. 10.4103/ejos.ejos_53_22.

Foster, P., Khaw, K.T., 2009. The eye: window to the soul or a mirror of systemic health? Heart. 95, 348–349. 10.1136/hrt.2008.158121.

Friedel, E.B.N., Beringer, M., Endres, D., Runge, K., Maier, S., Küchlin, S., Kornmeier, J., Domschke, K., Heinrich, S.P., Tebartz van Elst, L., Nickel, K., 2025. Optical coherence tomography in patients with major depressive disorder. BMC Psychiatry. 25, 356. 10.1186/s12888-025-06775-7.

Gao, X., Geng, T., Jiang, M., Huang, N., Zheng, Y., Belsky, D.W., Huang, T., 2023. Accelerated biological aging and risk of depression and anxiety: evidence from 424,299 UK Biobank participants. Nat Commun. 14, 2277. 10.1038/s41467-023-38013-7.

Gemelli, H., Fidalgo, T.M., Gracitelli, C.P.B., de Andrade, E.P., 2019. Retinal nerve fiber layer analysis in cocaine users. Psychiatry Res. 271, 226–229. 10.1016/j.psychres.2018.11.058.

Genc, A., Dalkıran, M., Pirdoğan Aydın, E., Türkyılmaz Uyar, E., Alkan, A., Guven, D., Özer Ö, A., Karamustafalıoğlu, O., 2019. The alteration of retinal nerve fibre layer thickness with repetitive transcranial magnetic stimulation in patients with treatment resistant major depression. Int J Psychiatry Clin Pract. 23, 57–61. 10.1080/13651501.2018.1480785.

Gonzalez-Diaz, J.M., Radua, J., Sanchez-Dalmau, B., Camos-Carreras, A., Zamora, D.C., Bernardo, M., 2022. Mapping Retinal Abnormalities in Psychosis: Meta-analytical Evidence for Focal Peripapillary and Macular Reductions. Schizophr Bull. 48, 1194–1205. 10.1093/schbul/sbac085.

Gooty, J., Banks, G., Loignon, A., Tonidandel, S., Williams, C., 2019. Meta-Analyses as a Multi-Level Model. Organizational Research Methods. 24, 109442811985747. 10.1177/1094428119857471.

Gupta, R., Advani, D., Yadav, D., Ambasta, R.K., Kumar, P., 2023. Dissecting the Relationship Between Neuropsychiatric and Neurodegenerative Disorders. Mol Neurobiol. 60, 6476–6529. 10.1007/s12035-023-03502-9.

Harwerth, R.S., Wheat, J.L., Rangaswamy, N.V., 2008. Age-related losses of retinal ganglion cells and axons. Invest Ophthalmol Vis Sci. 49, 4437–4443. 10.1167/iovs.08-1753.

Jayanthi, S., Daiwile, A.P., Cadet, J.L., 2021. Neurotoxicity of methamphetamine: Main effects and mechanisms. Exp Neurol. 344, 113795. 10.1016/j.expneurol.2021.113795.

Jung, K.I., Hong, S.Y., Shin, D., Lee, N.Y., Kim, T.S., Park, C.K., 2020. Attenuated Visual Function in Patients with Major Depressive Disorder. J. Clin. Med. 9, 10, 1951. 10.3390/jcm9061951.

Kalenderoglu, A., Egeli-Karatas, A., Hamdi Orum, M., Sevgi Karadag, A., 2025. Panic disorder causes an increase in choroid layer and a decrease in IPL, GCL, RNFL. Psychiatr Danub. 37, 38–45. 10.24869/psyd.2025.38.

Kalenderoglu, A., Orum, M.H., Karadag, A.S., Kustepe, A., Celik, M., Egilmez, O.B., Eken-Gedik, D., 2020. Increases in retinal nerve fiber layer thickness may represent the neuroprotective effect of cannabis: an optical coherence tomography study. J Addict Dis. 38, 280–290. 10.1080/10550887.2020.1754109.

Kallen, N.M., Cecere, G., Palpella, D., Rabe, F., Georgiadis, F., Badstübner, P., Edkins, V., Trindade, M., Homan, S., Omlor, W., Seifritz, E., Homan, P., 2024. The retina across the psychiatric spectrum: a systematic review and meta-analysis. medRxiv. 2024.2011.2007.24316925. 10.1101/2024.11.07.24316925.

Karadere, M.E., Sahin, T., Cobanoglu, E., Yildiz, V., 2020. Retinal Nerve Fiber Layer Thickness in Opioid Abusers. Psychiatry Clin. Psychopharmacology. 30, 369–373. 10.5455/PCP.20200702123911.

Kaufmann, T., van der Meer, D., Doan, N.T., Schwarz, E., Lund, M.J., Agartz, I., Alnæs, D., Barch, D.M., Baur-Streubel, R., Bertolino, A., Bettella, F., Beyer, M.K., Bøen, E., Borgwardt, S., Brandt, C.L., Buitelaar, J., Celius, E.G., Cervenka, S., Conzelmann, A., Córdova-Palomera, A., Dale, A.M., de Quervain, D.J.F., Di Carlo, P., Djurovic, S., Dørum, E.S., Eisenacher, S., Elvsåshagen, T., Espeseth, T., Fatouros-Bergman, H., Flyckt, L., Franke, B., Frei, O., Haatveit, B., Håberg, A.K., Harbo, H.F., Hartman, C.A., Heslenfeld, D., Hoekstra, P.J., Høgestøl, E.A., Jernigan, T.L., Jonassen, R., Jönsson, E.G., Farde, L., Flyckt, L., Engberg, G., Erhardt, S., Fatouros-Bergman, H., Cervenka, S., Schwieler, L., Piehl, F., Agartz, I., Collste, K., Victorsson, P., Malmqvist, A., Hedberg, M., Orhan, F., Kirsch, P., Kłoszewska, I., Kolskår, K.K., Landrø, N.I., Le Hellard, S., Lesch, K.-P., Lovestone, S., Lundervold, A., Lundervold, A.J., Maglanoc, L.A., Malt, U.F., Mecocci, P., Melle, I., Meyer-Lindenberg, A., Moberget, T., Norbom, L.B., Nordvik, J.E., Nyberg, L., Oosterlaan, J., Papalino, M., Papassotiropoulos, A., Pauli, P., Pergola, G., Persson, K., Richard, G., Rokicki, J., Sanders, A.-M., Selbæk, G., Shadrin, A.A., Smeland, O.B., Soininen, H., Sowa, P., Steen, V.M., Tsolaki, M., Ulrichsen, K.M., Vellas, B., Wang, L., Westman, E., Ziegler, G.C., Zink, M., Andreassen, O.A., Westlye, L.T., Karolinska Schizophrenia, P., 2019. Common brain disorders are associated with heritable patterns of apparent aging of the brain. Nature Neuroscience. 22, 1617–1623. 10.1038/s41593-019-0471-7.

Kaya, S., Kaya, M.K., 2023. OCT Findings in Patients with Methamphetamine Use Disorder. J. Pers. Med. 13, 12, 308. 10.3390/jpm13020308.

Kaya, Ş., Kaya, M.K., 2024. Assessment of RNFL and macular changes in the eye related to multiple substance use using OCT. Psychiatry Research: Neuroimaging. 345, 111889. 10.1016/j.pscychresns.2024.111889.

Khalil, M.A., Khalil, N.M., Esmael, A.F., El-Makawi, S.M., Saleh, A.A., Ayoub, D.R., 2023. Degenerative brain changes associated with tramadol use: an optical coherence tomography study. Middle East Curr. Psychiatry. 30, 100. 10.1186/s43045-023-00374-6.

Kim, H.M., Han, J.W., Park, Y.J., Bae, J.B., Woo, S.J., Kim, K.W., 2022. Association Between Retinal Layer Thickness and Cognitive Decline in Older Adults. JAMA Ophthalmology. 140, 683–690. 10.1001/jamaophthalmol.2022.1563.

Ko, F., Muthy, Z.A., Gallacher, J., Sudlow, C., Rees, G., Yang, Q., Keane, P.A., Petzold, A., Khaw, P.T., Reisman, C., Strouthidis, N.G., Foster, P.J., Patel, P.J., 2018. Association of Retinal Nerve Fiber Layer Thinning With Current and Future Cognitive Decline: A Study Using Optical Coherence Tomography. JAMA Neurol. 75, 1198–1205. 10.1001/jamaneurol.2018.1578.

Komatsu, H., Onoguchi, G., Jerotic, S., Kanahara, N., Kakuto, Y., Ono, T., Funakoshi, S., Yabana, T., Nakazawa, T., Tomita, H., 2022. Retinal layers and associated clinical factors in schizophrenia spectrum disorders: a systematic review and meta-analysis. Molecular psychiatry. 27, 3592–3616.

Leung, C.K.S., Yu, M., Weinreb, R.N., Ye, C., Liu, S., Lai, G., Lam, D.S.C., 2012. Retinal Nerve Fiber Layer Imaging with Spectral-Domain Optical Coherence Tomography: A Prospective Analysis of Age-Related Loss. Ophthalmology. 119, 731–737. 10.1016/j.ophtha.2011.10.010.

Liu, X., Lai, S.K., Ma, S.S., Yang, H., Liu, L., Yu, G.C., Zhong, S.M., Jia, Y.B., Zhong, J.X., 2022a. Development of a Novel Retina-Based Diagnostic Score for Early Detection of Major Depressive Disorder: An Interdisciplinary View. Front. Psychiatry. 13, 9, 897759. 10.3389/fpsyt.2022.897759.

Liu, Y., Huang, L., Wang, Z., Chen, J., Bian, Q., Sun, J., Jiang, L., Yang, F., 2021a. The changes in retinal nerve fiber layer and macular thickness in Chinese patients with alcohol dependency. Drug Alcohol Depend. 229, 109130. 10.1016/j.drugalcdep.2021.109130.

Liu, Y.H., Chen, J.X., Huang, L., Yan, S.X., Gao, D.F., Yang, F.D., 2022b. Association between changes in the retina with major depressive disorder and sleep quality. Journal of Affective Disorders. 311, 548–553. 10.1016/j.jad.2022.05.074.

Liu, Y.H., Tong, Y.S., Huang, Z., Chen, J.X., Yan, S.X., Yang, F.D., 2021b. Factors related to retinal nerve fiber layer thickness in bipolar disorder patients and major depression patients. BMC Psychiatry. 21, 10, 301. 10.1186/s12888-021-03270-7.

London, A., Benhar, I., Schwartz, M., 2013. The retina as a window to the brain-from eye research to CNS disorders. Nat Rev Neurol. 9, 44–53. 10.1038/nrneurol.2012.227.

Maes, M., Bonifacio, K.L., Morelli, N.R., Vargas, H.O., Moreira, E.G., St. Stoyanov, D., Barbosa, D.S., Carvalho, A.F., Nunes, S.O.V., 2018. Generalized Anxiety Disorder (GAD) and Comorbid Major Depression with GAD Are Characterized by Enhanced Nitro-oxidative Stress, Increased Lipid Peroxidation, and Lowered Lipid-Associated Antioxidant Defenses. Neurotoxicity Research. 34, 489–510. 10.1007/s12640-018-9906-2.

Malhi, G.S., Mann, J.J., 2018. Depression. The Lancet. 392, 2299–2312. 10.1016/S0140-6736(18)31948-2.

Moola S M.Z., Tufanaru C, Aromataris E, Sears K, Sfetcu R, Currie M, Qureshi R, Mattis P, Lisy K, Mu P-F, 2017. Sytematic reveiws of etiology and risk, In: Aromataris E, M.Z. (Ed.), Joanna Briggs Institute Reviewer’s Manual, / ed. /, The Joanna Briggs Institute, p. /.

Mutlu, U., Colijn, J.M., Ikram, M.A., Bonnemaijer, P.W.M., Licher, S., Wolters, F.J., Tiemeier, H., Koudstaal, P.J., Klaver, C.C.W., Ikram, M.K., 2018. Association of Retinal Neurodegeneration on Optical Coherence Tomography With Dementia: A Population-Based Study. JAMA Neurol. 75, 1256–1263. 10.1001/jamaneurol.2018.1563.

Mwanza, J.C., Durbin, M.K., Budenz, D.L., Girkin, C.A., Leung, C.K., Liebmann, J.M., Peace, J.H., Werner, J.S., Wollstein, G., 2011. Profile and predictors of normal ganglion cell-inner plexiform layer thickness measured with frequency-domain optical coherence tomography. Invest Ophthalmol Vis Sci. 52, 7872–7879. 10.1167/iovs.11-7896.

Nasir, E., ul Haq, I., Asghar, A., Nazir, T., Obaid, N., 2025. A Correlation Between Retinal Nerve Fibre Layer Thickness And Axial Length In Non-Glaucomatous Myopes. Journal of Rawalpindi Medical College. 29.

Nishida, T., Moghimi, S., Chang, A.C., Walker, E., Liebmann, J.M., Fazio, M.A., Girkin, C.A., Zangwill, L.M., Weinreb, R.N., 2022. Association of intraocular pressure with retinal nerve fiber layer thinning in patients with glaucoma. JAMA ophthalmology. 140, 1209–1216.

Noah, A.M., Almghairbi, D., Moppett, I.K., 2020. Optical coherence tomography in mild cognitive impairment - Systematic review and meta-analysis. Clin Neurol Neurosurg. 196, 106036. 10.1016/j.clineuro.2020.106036.

Orum, M.H., Kalenderoglu, A., 2020. Decreases in retinal nerve fiber layer thickness correlates with cumulative alcohol intake. J Addict Dis. 38, 400–410. 10.1080/10550887.2020.1776083.

Orum, M.H., Kalenderoglu, A., 2021. Acute opioid use may cause choroidal thinning and retinal nerve fiber layer increase. J Addict Dis. 39, 322–330. 10.1080/10550887.2021.1874816.

Orum, M.H., Kalenderoglu, A., Karadag, A.S., Hocaoglu, C., 2022. Retinal nerve fiber layer decrease and choroidal layer increase after four weeks of buprenorphine/naloxone maintenance treatment in opioid use disorder. Eur. J. Psychiatry. 36, 260–270. 10.1016/j.ejpsy.2022.04.001.

Ozisik, G.G., Kiraz, S., 2023. Evaluation of retinal thickness measured by optical coherence tomography in patients with generalized anxiety disorder. Photodiagnosis Photodyn Ther. 44, 103766. 10.1016/j.pdpdt.2023.103766.

Özsoy, F., Kulu, M., Özarslan, Y., Korkmaz, S., 2023. Relationship between optical coherence tomography findings and clinical variables in patients with opiate use disorder. Arq Bras Oftalmol. 86, 20–26. 10.5935/0004-2749.20230013.

Petzold, A., Chua, S.Y.L., Khawaja, A.P., Keane, P.A., Khaw, P.T., Reisman, C., Dhillon, B., Strouthidis, N.G., Foster, P.J., Patel, P.J., 2021. Retinal asymmetry in multiple sclerosis. Brain. 144, 224–235. 10.1093/brain/awaa361.

Petzold, A., Fraser, C.L., Abegg, M., Alroughani, R., Alshowaeir, D., Alvarenga, R., Andris, C., Asgari, N., Barnett, Y., Battistella, R., Behbehani, R., Berger, T., Bikbov, M.M., Biotti, D., Biousse, V., Boschi, A., Brazdil, M., Brezhnev, A., Calabresi, P.A., Cordonnier, M., Costello, F., Cruz, F.M., Cunha, L.P., Daoudi, S., Deschamps, R., de Seze, J., Diem, R., Etemadifar, M., Flores-Rivera, J., Fonseca, P., Frederiksen, J., Frohman, E., Frohman, T., Tilikete, C.F., Fujihara, K., Gálvez, A., Gouider, R., Gracia, F., Grigoriadis, N., Guajardo, J.M., Habek, M., Hawlina, M., Martínez-Lapiscina, E.H., Hooker, J., Hor, J.Y., Howlett, W., Huang-Link, Y., Idrissova, Z., Illes, Z., Jancic, J., Jindahra, P., Karussis, D., Kerty, E., Kim, H.J., Lagrèze, W., Leocani, L., Levin, N., Liskova, P., Liu, Y., Maiga, Y., Marignier, R., McGuigan, C., Meira, D., Merle, H., Monteiro, M.L.R., Moodley, A., Moura, F., Muñoz, S., Mustafa, S., Nakashima, I., Noval, S., Oehninger, C., Ogun, O., Omoti, A., Pandit, L., Paul, F., Rebolleda, G., Reddel, S., Rejdak, K., Rejdak, R., Rodriguez-Morales, A.J., Rougier, M.B., Sa, M.J., Sanchez-Dalmau, B., Saylor, D., Shatriah, I., Siva, A., Stiebel-Kalish, H., Szatmary, G., Ta, L., Tenembaum, S., Tran, H., Trufanov, Y., van Pesch, V., Wang, A.G., Wattjes, M.P., Willoughby, E., Zakaria, M., Zvornicanin, J., Balcer, L., Plant, G.T., 2022. Diagnosis and classification of optic neuritis. Lancet Neurol. 21, 1120–1134. 10.1016/s1474-4422(22)00200-9.

Prasannakumar, A., Kumar, V., Mailankody, P., Appaji, A., Battu, R., Berendschot, T.T., Rao, N.P., 2023. A systematic review and meta-analysis of optical coherence tomography studies in schizophrenia, bipolar disorder and major depressive disorder. The World Journal of Biological Psychiatry. 24, 707–720.

Ruggeri, F., Fumi, D., Bassis, L., Pippo, M.D., Abdolrahimzadeh, S., 2025. The Role of the Ganglion Cell Layer as an OCT Biomarker in Neurodegenerative Diseases. J Integr Neurosci. 24, 26039. 10.31083/jin26039.

Scheffler, F., Ipser, J., Pancholi, D., Murphy, A., Cao, Z., Ottino-González, J., Thompson, P.M., Shoptaw, S., Conrod, P., Mackey, S., Garavan, H., Stein, D.J., 2024. Mega-analysis of the brain-age gap in substance use disorder: An ENIGMA Addiction working group study. Addiction. 119, 1937–1946. 10.1111/add.16621.

Schönfeldt-Lecuona, C., Schmidt, A., Kregel, T., Kassubek, J., Dreyhaupt, J., Freudenmann, R.W., Connemann, B.J., Pinkhardt, E.H., Gahr, M., 2018. Retinal changes in patients with major depressive disorder - A controlled optical coherence tomography study. J Affect Disord. 227, 665–671. 10.1016/j.jad.2017.11.077.

Sönmez, I., Kösger, F., Aykan, Ü., 2017. Retinal Nerve Fiber Layer Thickness Measurement by Spectral-Domain Optical Coherence Tomography in Patients with Major Depressive Disorder. Noropsikiyatri Ars. 54, 62–66. 10.5152/npa.2015.10115.

Steel, Z., Marnane, C., Iranpour, C., Chey, T., Jackson, J.W., Patel, V., Silove, D., 2014. The global prevalence of common mental disorders: a systematic review and meta-analysis 1980-2013. Int J Epidemiol. 43, 476–493. 10.1093/ije/dyu038.

Stuart, K.V., Luben, R.N., Warwick, A.N., Madjedi, K.M., Patel, P.J., Biradar, M.I., Sun, Z., Chia, M.A., Pasquale, L.R., Wiggs, J.L., Kang, J.H., Kim, J., Aschard, H., Tran, J.H., Lentjes, M.A.H., Foster, P.J., Khawaja, A.P., 2023. The Association of Alcohol Consumption with Glaucoma and Related Traits: Findings from the UK Biobank. Ophthalmol Glaucoma. 6, 366–379. 10.1016/j.ogla.2022.11.008.

Sung, K.R., Wollstein, G., Bilonick, R.A., Townsend, K.A., Ishikawa, H., Kagemann, L., Noecker, R.J., Fujimoto, J.G., Schuman, J.S., 2009. Effects of age on optical coherence tomography measurements of healthy retinal nerve fiber layer, macula, and optic nerve head. Ophthalmology. 116, 1119–1124.

Talebnejad, M.R., Khazaei, P., Jahanbani-Ardakani, H., Saberikia, Z., Moghimi Sarani, E., Khalili, M.R., 2020. Effects of chronic methamphetamine abuse on the retinal nerve fiber layer, ganglion cell layer and Bruch’s membrane opening minimum rim width. Neurotoxicology. 80, 140–143. 10.1016/j.neuro.2020.07.005.

Thomson, K.L., Yeo, J.M., Waddell, B., Cameron, J.R., Pal, S., 2015. A systematic review and meta-analysis of retinal nerve fiber layer change in dementia, using optical coherence tomography. Alzheimer’s & Dementia: Diagnosis, Assessment & Disease Monitoring. 1, 136–143.

Turhan, N.O., Arisoy, O., Ulas, F., Bugdayci, G., Guelner, M.A., 2024. Vitamin D: An Overlooked Parameter in Studies of Depression Using Optic Coherence Tomography. Noropsikiyatri Ars. 61, 66–72. 10.29399/npa.28369.

Vermani, M., Marcus, M., Katzman, M.A., 2011. Rates of detection of mood and anxiety disorders in primary care: a descriptive, cross-sectional study. Prim Care Companion CNS Disord. 13. 10.4088/PCC.10m01013.

Viechtbauer, W., 2010. Conducting Meta-Analyses in R with the metafor Package. Journal of Statistical Software. 36, 1–48. 10.18637/jss.v036.i03.

Volkow, N.D., Boyle, M., 2018. Neuroscience of Addiction: Relevance to Prevention and Treatment. Am J Psychiatry. 175, 729–740. 10.1176/appi.ajp.2018.17101174.

Wagner, S.K., Romero-Bascones, D., Cortina-Borja, M., Williamson, D.J., Struyven, R.R., Zhou, Y., Patel, S., Weil, R.S., Antoniades, C.A., Topol, E.J., Korot, E., Foster, P.J., Balaskas, K., Ayala, U., Barrenechea, M., Gabilondo, I., Schapira, A.H.V., Khawaja, A.P., Patel, P.J., Rahi, J.S., Denniston, A.K., Petzold, A., Keane, P.A., 2023. Retinal Optical Coherence Tomography Features Associated With Incident and Prevalent Parkinson Disease. Neurology. 101, e1581–e1593. 10.1212/wnl.0000000000207727.

Wang, Y., Li, C., Liu, L., Yang, Y., He, X., Li, G., Zheng, X.Z., Ren, Y., Zhao, H.P., Du, Z.C., Jiang, J.R., Kuang, Y., Jia, F.J., Yu, H.H., Yang, X.H., 2024. Association of Retinal Neurovascular Impairment with Disease Severity in Patients with Major Depressive Disorder: An Optical Coherence Tomography Angiography Study. Psychol. Res. Behav. Manag. 17, 1573–1585. 10.2147/prbm.S443146.

World Health Organization, 2025. World mental health today: latest data Geneva.

Wrigglesworth, J., Ward, P., Harding, I.H., Nilaweera, D., Wu, Z., Woods, R.L., Ryan, J., 2021. Factors associated with brain ageing-a systematic review. BMC neurology. 21, 312.

Xiao, Q., Li, F.L., Jiang, F.R., Zhang, Z.J., Xu, B., 2024. The prospects for early detection with optical coherence tomography (OCT) and OCT angiography in major depressive disorder. Journal of Affective Disorders. 347, 8–14. 10.1016/j.jad.2023.11.031.

Yildiz, M., Alim, S., Batmaz, S., Demir, S., Songur, E., Ortak, H., Demirci, K., 2016. Duration of the depressive episode is correlated with ganglion cell inner plexifrom layer and nasal retinal fiber layer thicknesses: Optical coherence tomography findings in major depression. Psychiatry Res. Neuroimaging. 251, 60–66. 10.1016/j.pscychresns.2016.04.011.

Youm, D.J., Kim, H., Shim, S.H., Jang, H.J., Kim, J.M., Park, K.H., Choi, C.Y., Cho, J.G., 2012. The effect of various factors on variability of retinal nerve fiber layer thickness measurements using optical coherence tomography. Korean Journal of Ophthalmology. 26, 104–110.

Zeppieri, M., Marsili, S., Enaholo, E.S., Shuaibu, A.O., Uwagboe, N., Salati, C., Spadea, L., Musa, M., 2023. Optical Coherence Tomography (OCT): A Brief Look at the Uses and Technological Evolution of Ophthalmology. Medicina (Kaunas). 59. 10.3390/medicina59122114.

Zhang, Z., Chen, X., Wu, S., Chen, X., Wang, X., Liu, C., Zeng, N., Liu, Y., Huo, T., Liu, X., Shi, L., Yan, W., Yuan, K., Meng, S., Wu, P., Javed, A., Alibudbud, R., Shi, J., Sun, Y., Lu, L., Bao, Y., 2026. Global, regional and national burden of anxiety and depression disorders from 1990 to 2021, and forecasts up to 2040. Journal of Affective Disorders. 393, 120299. 10.1016/j.jad.2025.120299.

Zhu, Z., Cheung, H., Ding, K., Tan, Q., Zhu, Y., Cheng, D., Kong, S., Yin, X., Wang, D., Zhuang, K., Chen, Y., Zhou, J., Luo, Y., Li, Z., Chen, L., 2026. Biological age acceleration associated with mental and behavioural disorders: Evidence from the UK biobank cohort. J Affect Disord. 401, 121265. 10.1016/j.jad.2026.121265.

