## Supplementary Materials for "Retinal Thickness in Anxiety, Depression, and Substance Use Disorders: A Systematic Review and Meta-Analysis of Optical Coherence Tomography (OCT) Studies"

Supplementary Table S1: Search Strategy

**09/07/2024**

| **PubMed** |  |  |  |  |  |  |  |  |
| --- | --- | --- | --- | --- | --- | --- | --- | --- |
| ("Tomography, Optical Coherence"[Mesh]) AND "Substance-Related Disorders"[Mesh] | | | | | | | | 66 |
| ("Tomography, Optical Coherence"[Mesh]) AND ( "Anxiety"[Mesh] OR "Anxiety Disorders"[Mesh] ) | | | | | | | | 17 |
| ("Tomography, Optical Coherence"[Mesh]) AND ( "Depression"[Mesh] OR "Depressive Disorder"[Mesh] ) | | | | | | | | 30 |
| **Scopus** |  |  |  |  |  |  |  |  |
| TITLE-ABS-KEY ( "optical coherence tomography" ) AND TITLE-ABS-KEY ( "substance use disorder*" OR "Substance use" OR "substance-related disorder*" ) ) | | | | | | | | 26 |
| (TITLE-ABS-KEY ( "tomography, optical coherence" ) AND TITLE-ABS-KEY ( {anxiety} ) OR TITLE-ABS-KEY ( "anxiety related disorder*" ) ) | | | | | | | | 69 |
| (TITLE-ABS-KEY ( "tomography, optical coherence" ) AND TITLE-ABS-KEY ( {depression} ) OR TITLE-ABS-KEY ( "depressive disorder*" ) ) | | | | | | | | 329 |
| **Web of Science** |  |  |  |  |  |  |  |  |
| ((ALL=(optical coherence tomography)) AND ALL=(substance use disorder)) | | | | | | | | 15 |
| ((ALL=(optical coherence tomography)) AND ALL=(anxiety disorder)) | | | | | | | | 18 |
| ((ALL=(optical coherence tomography)) AND (ALL=(depress* disorder))) | | | | | | | | 76 |
| **MedRxiv/BioXRiv** | Limited to subject: psychiatry and clinical psychology | | | | | | | |
| "optical coherence tomography" AND "substance use" OR "substance related disorder" | | | | | | | | 71 |
| "optical coherence tomography" AND "anxiety related disorder" | | | | | | | | 105 |
| "optical coherence tomography" AND "depression" OR "depressive disorder" | | | | | | | | 134 |
| **All Trials** |  |  |  |  |  |  |  |  |
| / |  |  |  |  |  |  |  |  |
| **Abandoned Trials** |  |  |  |  |  |  |  |  |
| / |  |  |  |  |  |  |  |  |
| **09/08/2024** |  |  |  |  |  |  |  |  |
| **Clinical Trials.gov** |  |  |  |  |  |  |  |  |
| Condition: Depression; Interventions: Optical Coherence Tomography/OCT | | | | | | | | 2 |
| Condition: Anxiety; Interventions: Optical Coherence Tomography/OCT | | | | | | | | 0 |
| Condition: Substance Use Disorders/SUD; Interventions: Optical Coherence Tomography/OCT | | | | | | | | 0 |

**12/01/2026**

| **PubMed** |  |  |  | |
| --- | --- | --- | --- | --- |
| ("Tomography, Optical Coherence"[Mesh]) AND "Substance-Related Disorders"[Mesh] | | | | 1 |
| ("Tomography, Optical Coherence"[Mesh]) AND ( "Anxiety"[Mesh] OR "Anxiety Disorders"[Mesh] ) | | | | 1 |
| ("Tomography, Optical Coherence"[Mesh]) AND ( "Depression"[Mesh] OR "Depressive Disorder"[Mesh] ) | | | | 1 |

Supplement Table S2 - Publication Bias (Funnel Plot Asymmetry)

| Analysis / Model | Predictor | Estimate | Standard Error | z-value | p-value | 95% CI Lower | 95% CI Upper | Residual Heterogeneity | Test of Moderators |
| --- | --- | --- | --- | --- | --- | --- | --- | --- | --- |
| **Model (Overall without covariates)** | Intercept | 1.5741 | 0.5719 | 2.4026 | 0.0163 | 0.2532 | 2.4959 | QE(df = 143) = 727.0348, p < .0001 | QM(df = 1) = 8.5835, p-val = 0.00431 |
|  | sei | -6.2583 | 2.1911 | -2.8571 | 0.0043 | -10.547 | -1.9691 |  |  |
| **Model (Overall with covariates)** | Intercept | 1.3664 | 0.6713 | 2.0345 | 0.0418 | 0.0507 | 2.6822 | QE(df = 131) = 709.9109, p < .0001 | QM(df = 3) = 9.2270, p-val = 0.0264 |
|  | sei | -6.4965 | 2.4188 | -2.6859 | 0.0072 | -11.2372 | -1.7558 |  |  |
|  | Mean Age | 0.0032 | 0.0838 | 0.2830 | 0.4077 | -0.0043 | 0.0108 |  |  |
|  | Mean Sex (% Male) | -0.2604 | 0.5967 | -0.4364 | 0.6626 | -1.4300 | 0.9092 |  |  |
| **Sensitivity (no Orum 2022)** | Intercept | 1.6940 | 0.5567 | 2.8438 | 0.0045 | 0.5265 | 2.8615 | QE(df = 127) = 668.8383, p < .0001 | QM(df = 1) = 11.1466, p-val = 0.0008 |
|  | sei | -7.5476 | 2.2698 | -3.3308 | 0.0008 | -11.9789 | -3.1169 |  |  |
| **Sensitivity (no outliers)** | Intercept | 0.3822 | 0.3870 | 0.9874 | 0.3234 | -0.3764 | 1.1407 | QE(df = 126) = 539.1431, p-val < .0001 | QM(df = 1) = 1.3726, p-val = 0.2414 |
|  | sei | -1.8072 | 1.5425 | -1.1716 | 0.2414 | -4.8305 | 1.2161 |  |  |
| **Anxiety (CMT)** | Intercept | 0.4702 | -- | -- | -- | -1.6210 | 2.5614 | -- | -- |
|  | sei | -- | -- | -0.5031 | 0.6149 | -- | -- |  |  |
| **Depression (Overall)** | Intercept | -1.2422 | 0.3757 | -3.3067 | 0.0009 | -1.9785 | -0.5059 | QE(df = 25) = 35.3345, p-val = 0.0823 | QM(df = 1) = 10.6161, p-val = 0.0011 |
|  | sei | 5.5903 | 1.7157 | 3.2582 | 0.0011 | 2.2275 | 8.9530 |  |  |
| **Depression (Superior RNFL)** | Intercept | -0.3694 | 1.8161 | -0.2034 | 0.8389 | -3.9288 | 3.1905 | QE(df = 2) = 0.9448, p-val = 0.6235 | QM(df = 1) = 0.0128, p-val = 0.9100 |
|  | sei | 1.1035 | 9.7582 | 0.1131 | 0.9100 | -18.0224 | 20.2293 |  |  |
| **Depression (Inferior RNFL)** | Intercept | -0.9686 | 2.5680 | -0.3775 | 0.7058 | -5.9979 | 4.0607 | QE(df = 2) = 3.7831, p-val = 0.1508 | QM(df = 1) = 0.1093, p-val = 0.7409 |
|  | sei | 4.5434 | 13.7415 | 0.3306 | 0.7409 | -22.3894 | 31.4762 |  |  |
| **Depression (Nasal RNFL)** | Intercept | 2.5205 | 7.2640 | 0.3470 | 0.7286 | -11.7167 | 16.7578 | QE(df = 1) = 6.3188, p-val = 0.0119 | QM(df = 1) = 0.1293, p-val = 0.7192 |
|  | sei | -13.6137 | 37.8668 | -0.3595 | 0.7192 | -87.8313 | 60.6039 |  |  |
| **Depression (Temporal RNFL)** | Intercept | 2.2495 | 4.1358 | 0.5429 | 0.5855 | -5.8565 | 10.3556 | QE(df = 1) = 2.0929, p-val = 0.1480 | QM(df = 1) = 0.3299, p-val = 0.5657 |
|  | sei | -12.4263 | 21.6346 | -0.5744 | 0.5657 | -54.8294 | 29.9768 |  |  |
| **Depression (Global RNFL)** | Intercept | -1.2392 | 0.4409 | -3.0149 | 0.0026 | -2.1334 | -0.4651 | QE(df = 11) = 18.1753, p-val = 0.0776 | QM(df = 1) = 9.4131, p-val = 0.0022 |
|  | sei | 5.9160 | 1.9282 | 3.0681 | 0.0022 | 2.1367 | 9.6953 |  |  |
| **SUD (Overall)** | Intercept | 1.8123 | 0.7789 | 2.3267 | 0.0200 | 0.2856 | 3.3390 | QE(df = 113) = 628.8015, p < .0001 | QM(df = 1) = 9.6864, p-val = 0.0019 |
|  | sei | -8.4122 | 2.7657 | -3.0397 | 0.0019 | -13.7152 | -3.1092 |  |  |
| **SUD (CMT)** | Intercept | 4.1799 | 2.0565 | 2.0296 | 0.0424 | 0.1432 | 8.2046 | QE(df = 8) = 25.2458, p-val = 0.0014 | QM(df = 1) = 5.3302, p-val = 0.0210 |
|  | sei | -17.1715 | 7.4376 | -2.3087 | 0.0210 | -31.7490 | -2.5940 |  |  |
| **SUD (GCL)** | Intercept | 0.5386 | 0.4558 | 1.1772 | 0.2391 | -0.3568 | 1.4301 | QE(df = 4) = 4.8079, p-val = 0.2989 | QM(df = 1) = 1.3199, p-val = 0.2506 |
|  | sei | -2.4907 | 2.1680 | -1.1489 | 0.2506 | -6.7398 | 1.7584 |  |  |
| **SUD (Superior RNFL)** | Intercept | 0.0232 | 1.1852 | 0.0196 | 0.9844 | -2.2998 | 2.3462 | QE(df = 22) = 95.8966, p < .0001 | QM(df = 1) = 0.0002, p-val = 0.9888 |
|  | sei | 0.0663 | 4.7389 | 0.0140 | 0.9888 | -9.2208 | 9.3563 |  |  |
| **SUD (Inferior RNFL)** | Intercept | 0.5148 | 0.8424 | 0.6111 | 0.5412 | -1.1363 | 2.1659 | QE(df = 22) = 49.8833, p-val = 0.0008 | QM(df = 1) = 0.3887, p-val = 0.5330 |
|  | sei | -2.1315 | 3.4187 | -0.6235 | 0.5330 | -8.8319 | 4.5689 |  |  |
| **SUD (Nasal RNFL)** | Intercept | -0.0871 | 1.0053 | -0.0866 | 0.9310 | -2.0575 | 1.8833 | QE(df = 14) = 44.6727, p < .0001 | QM(df = 1) = 0.0022, p-val = 0.9627 |
|  | sei | 0.1394 | 4.1184 | 0.0468 | 0.9627 | -7.919 | 8.3007 |  |  |
| **SUD (Temporal RNFL)** | Intercept | -0.1922 | 1.3133 | -0.1225 | 0.9017 | -2.7382 | 2.4118 | QE(df = 14) = 84.3086, p < .0001 | QM(df = 1) = 0.0253, p-val = 0.8735 |
|  | sei | 0.8413 | 5.2655 | 0.1592 | 0.8735 | -9.5181 | 11.2007 |  |  |
| **SUD (Global RNFL)** | Intercept | 1.8638 | 1.8038 | -0.4510 | 0.5613 | -6.6528 | 2.5247 | QE(df = 17) = 257.2961, p < .0001 | QM(df = 1) = 0.1214, p-val = 0.7275 |
|  | sei | 2.2411 | 8.4313 | 0.3455 | 0.7275 | -10.3640 | 14.8402 |  |  |

Supplementary Figures S1: Detailed Forest Plot - Overall Comparison

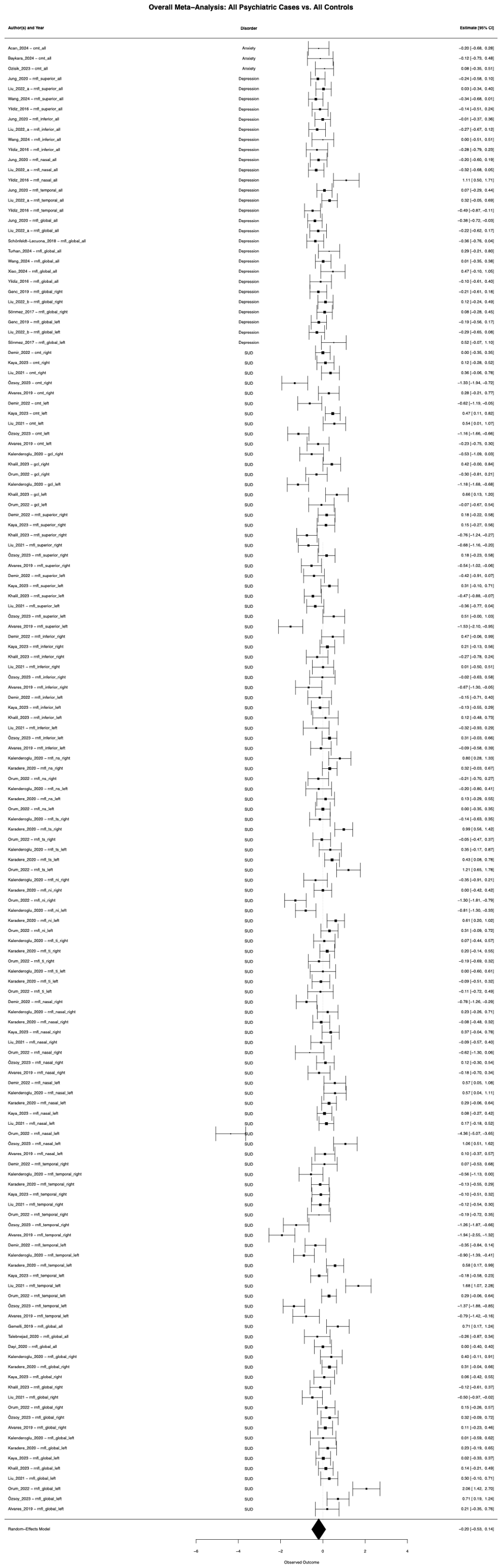

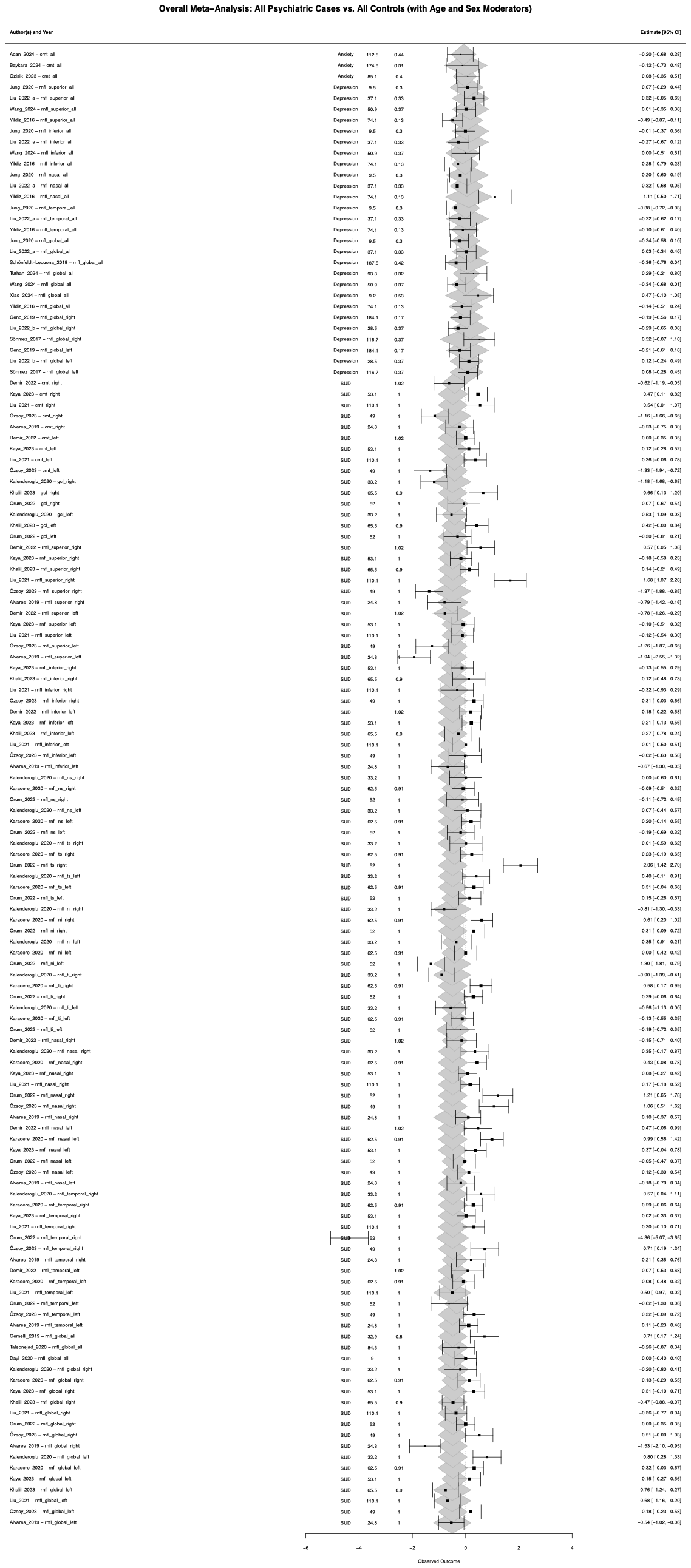

Supplementary Figure S2: Forest Plots - Depression subgroups of the RNFL

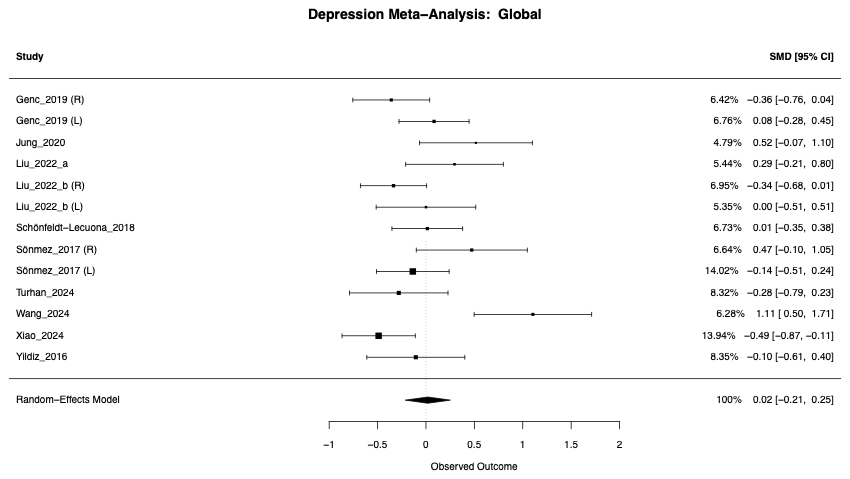

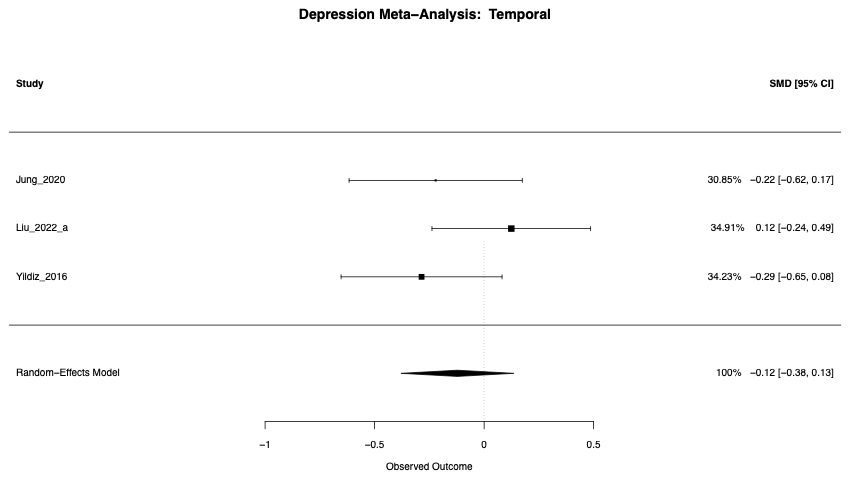

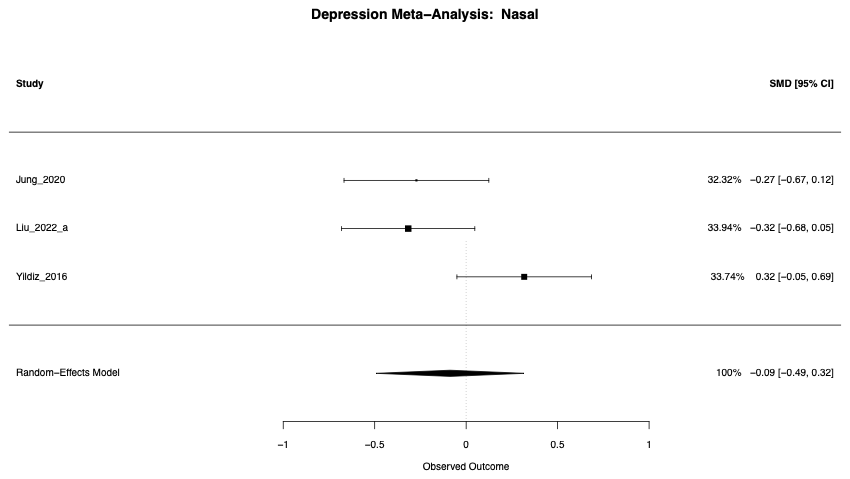

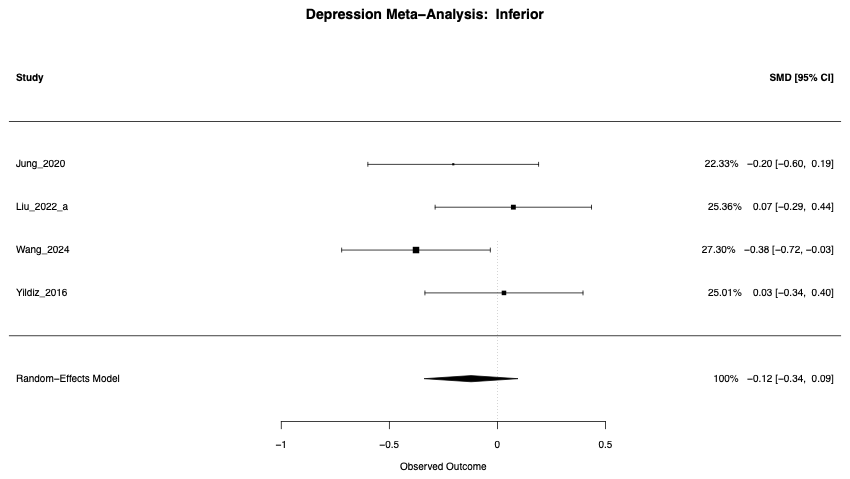

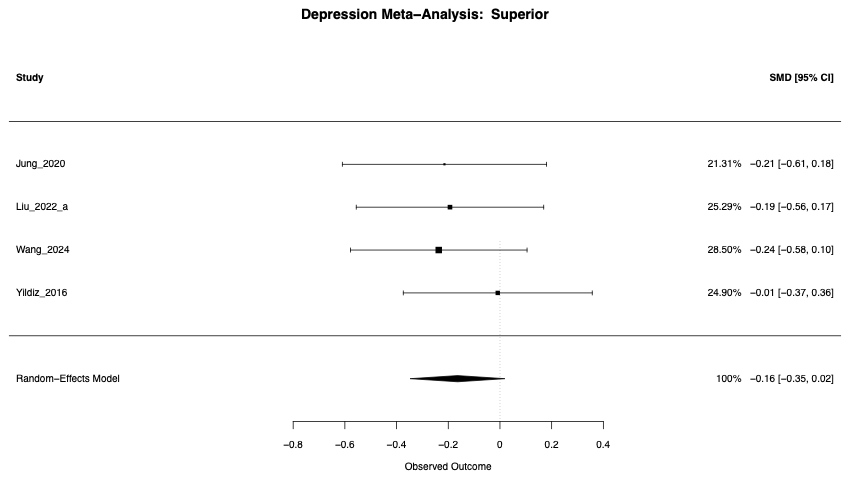

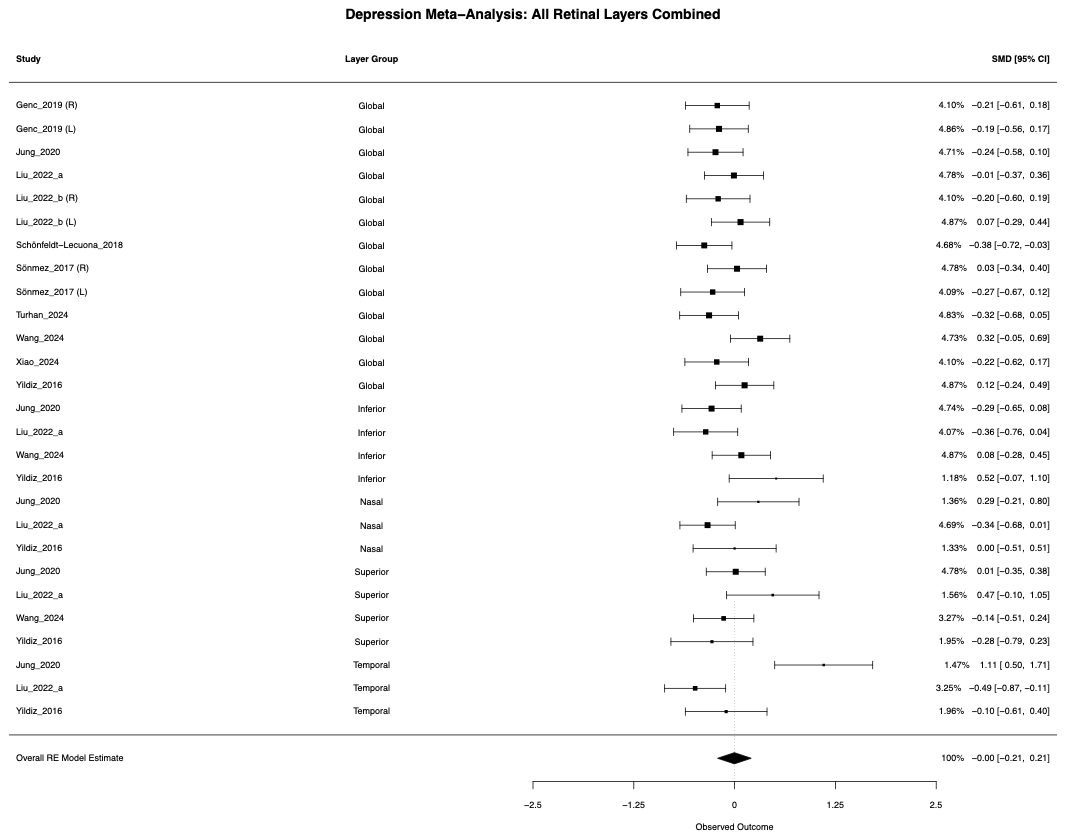

Supplementary Figure S3: Forest plots - SUD Layer Analysis

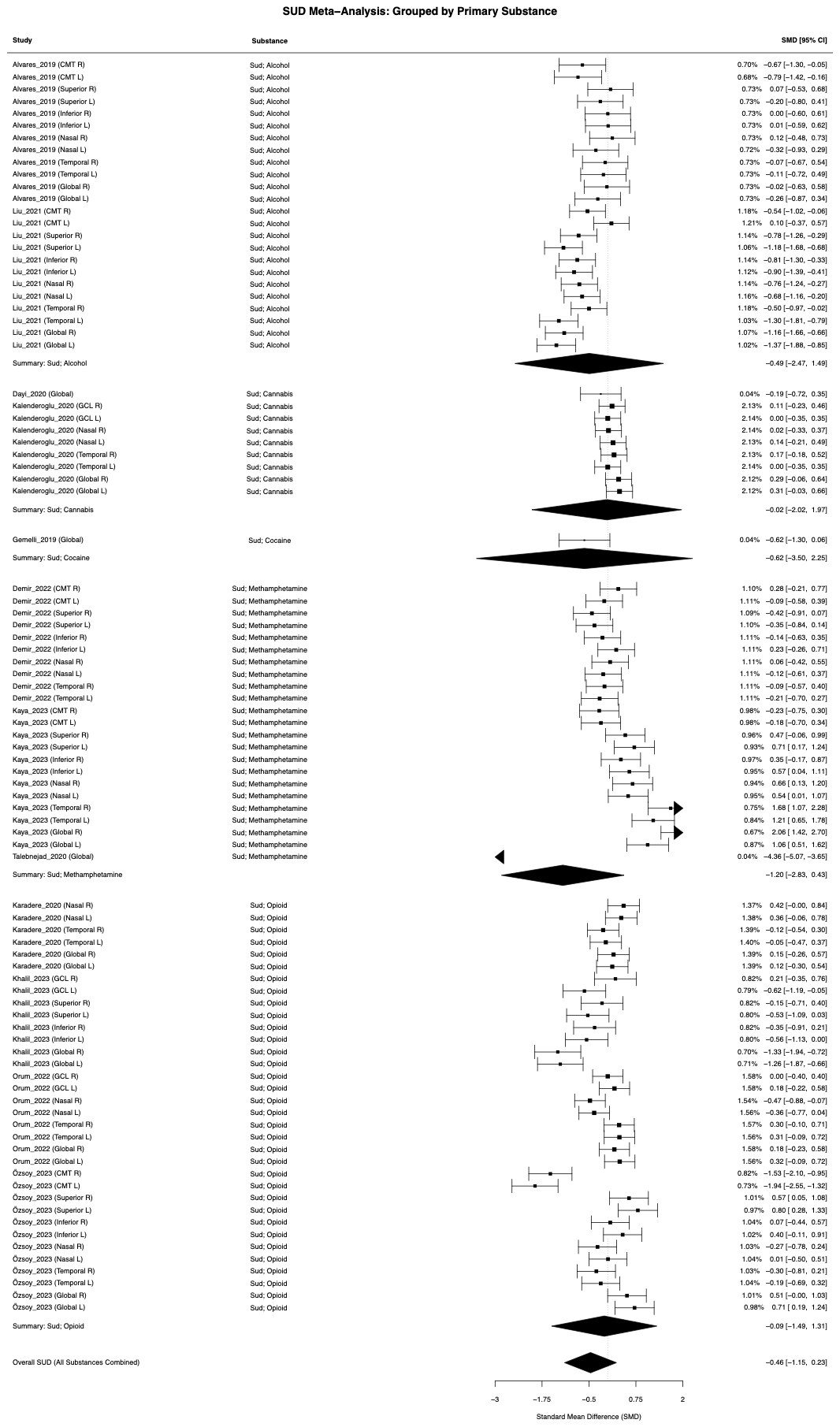

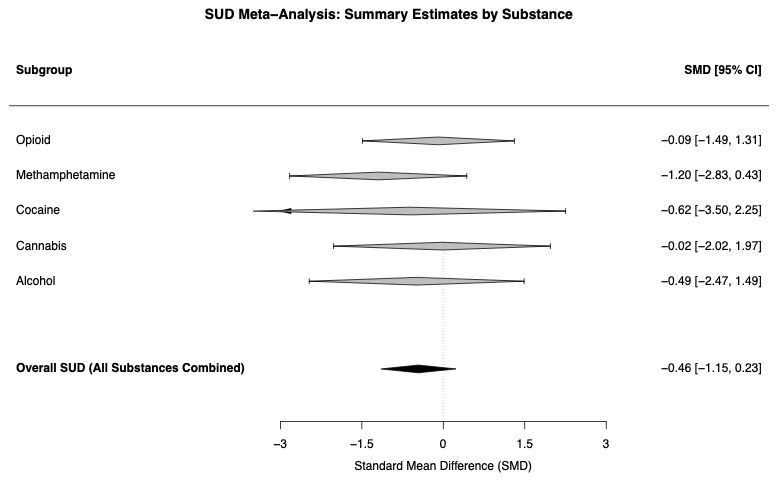

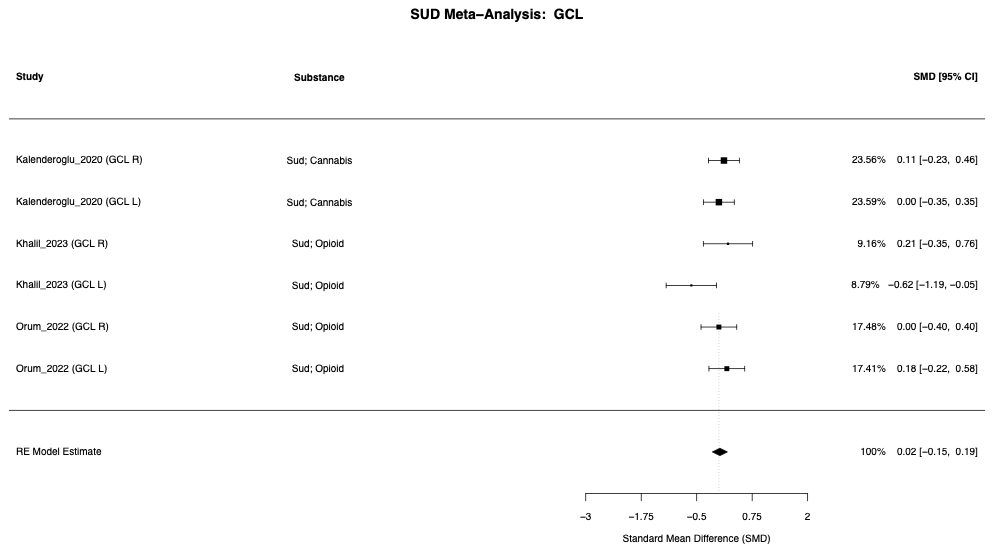

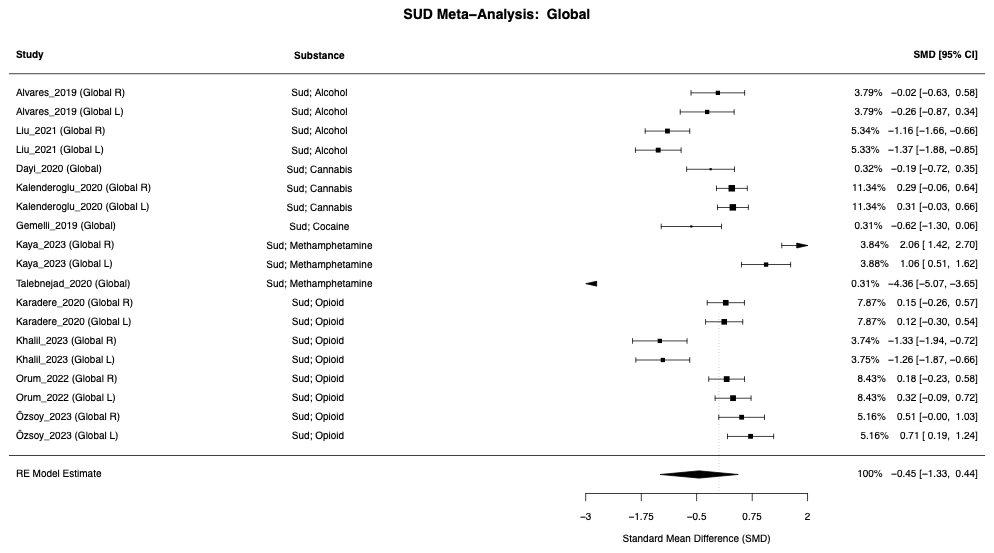

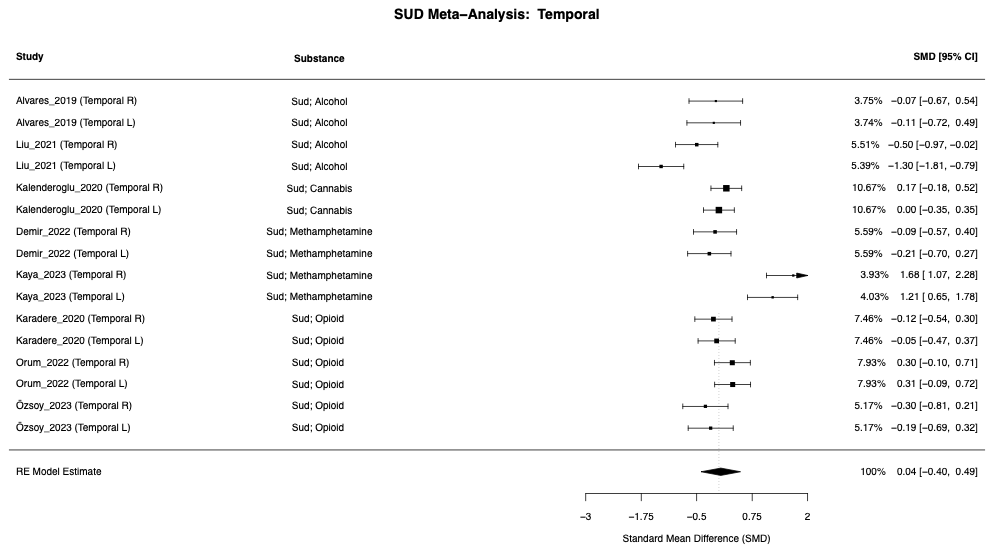

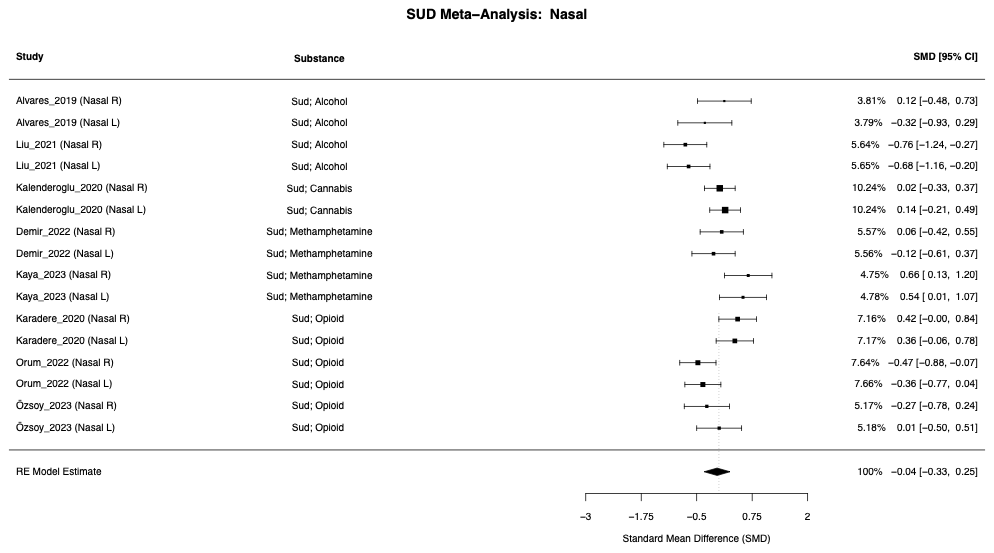

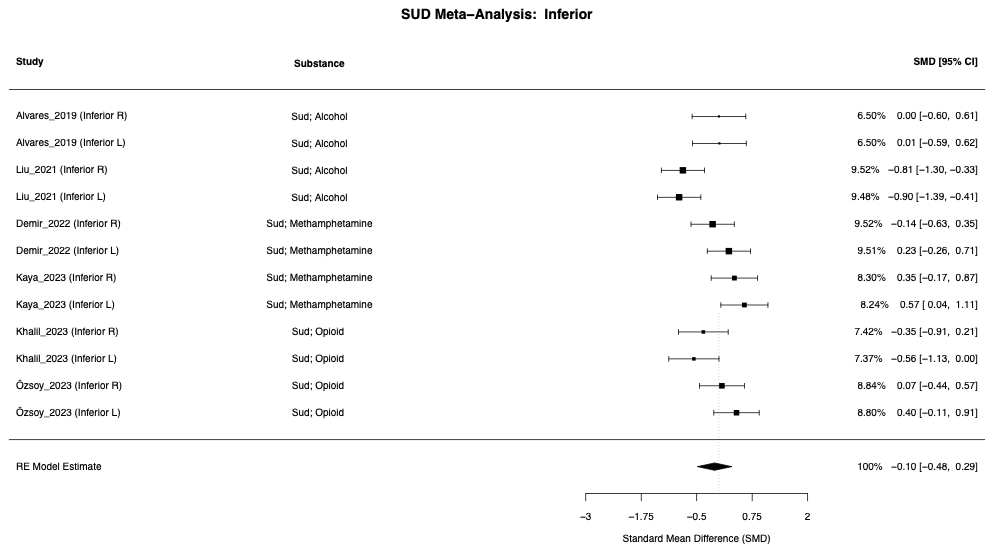

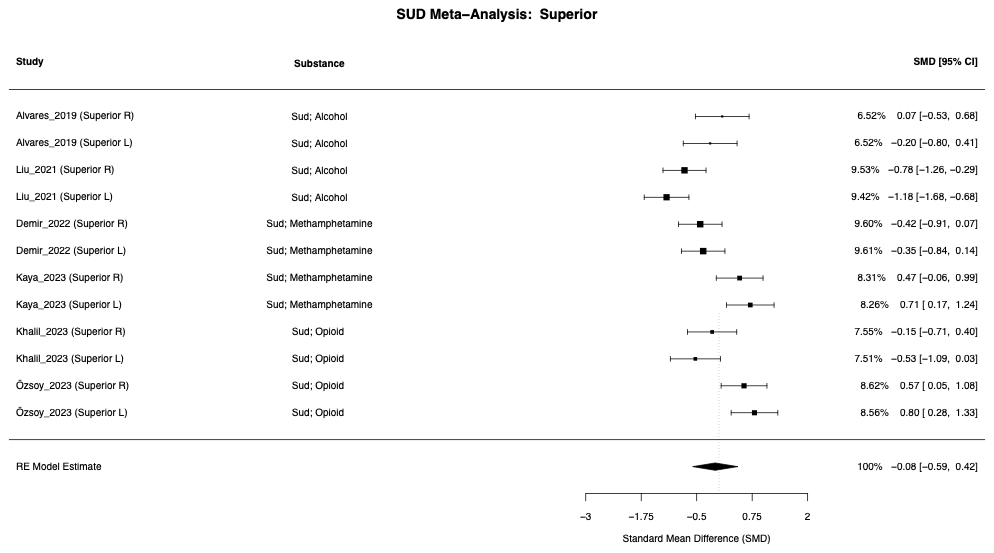

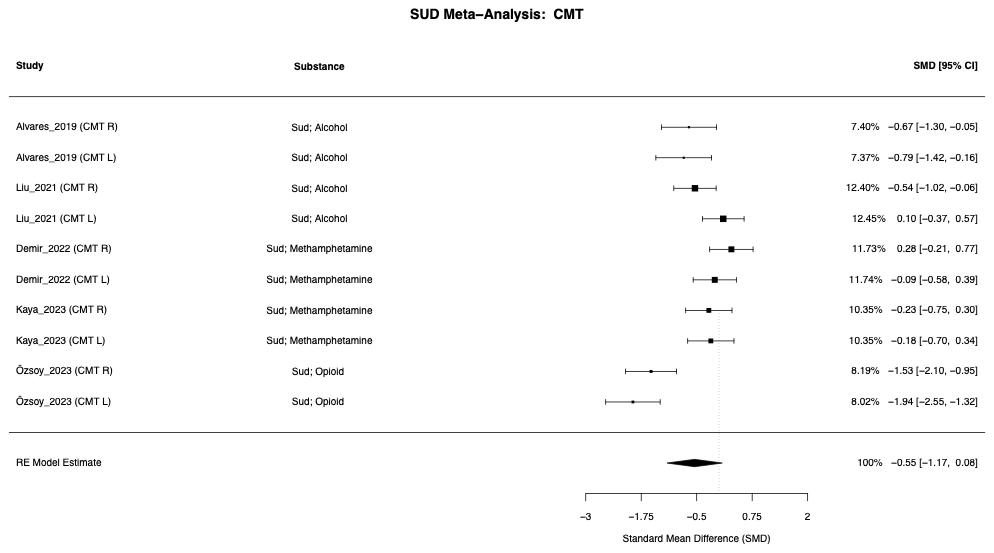

Forest Plots: Sensitivity Analysis

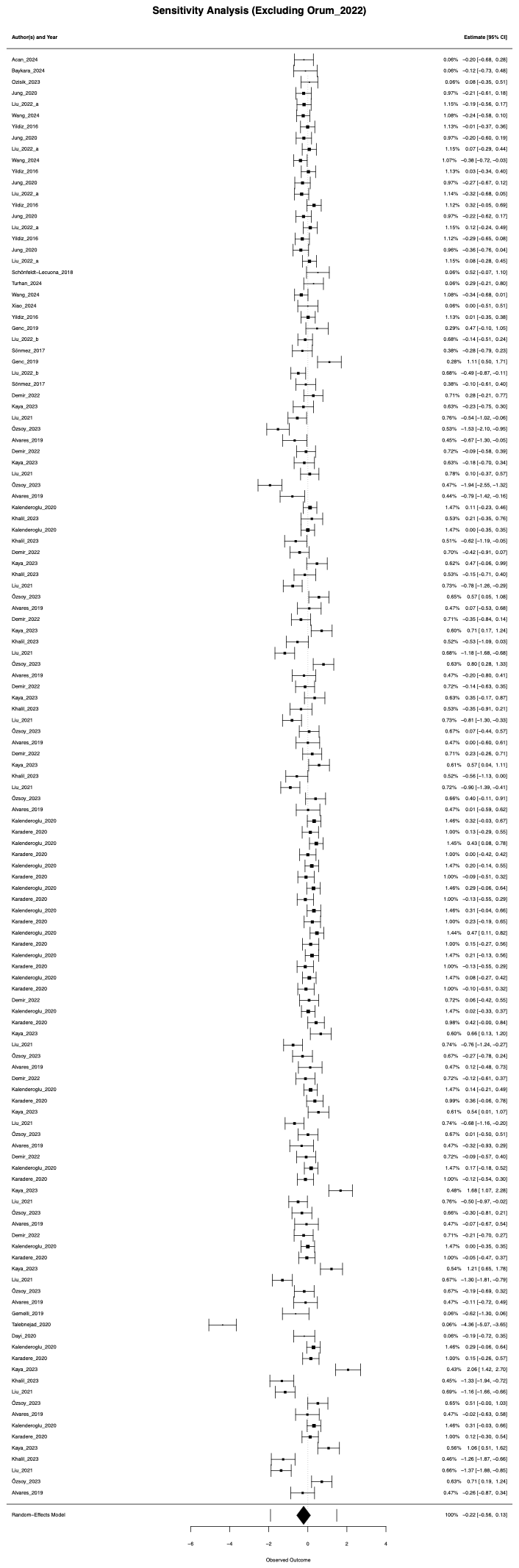

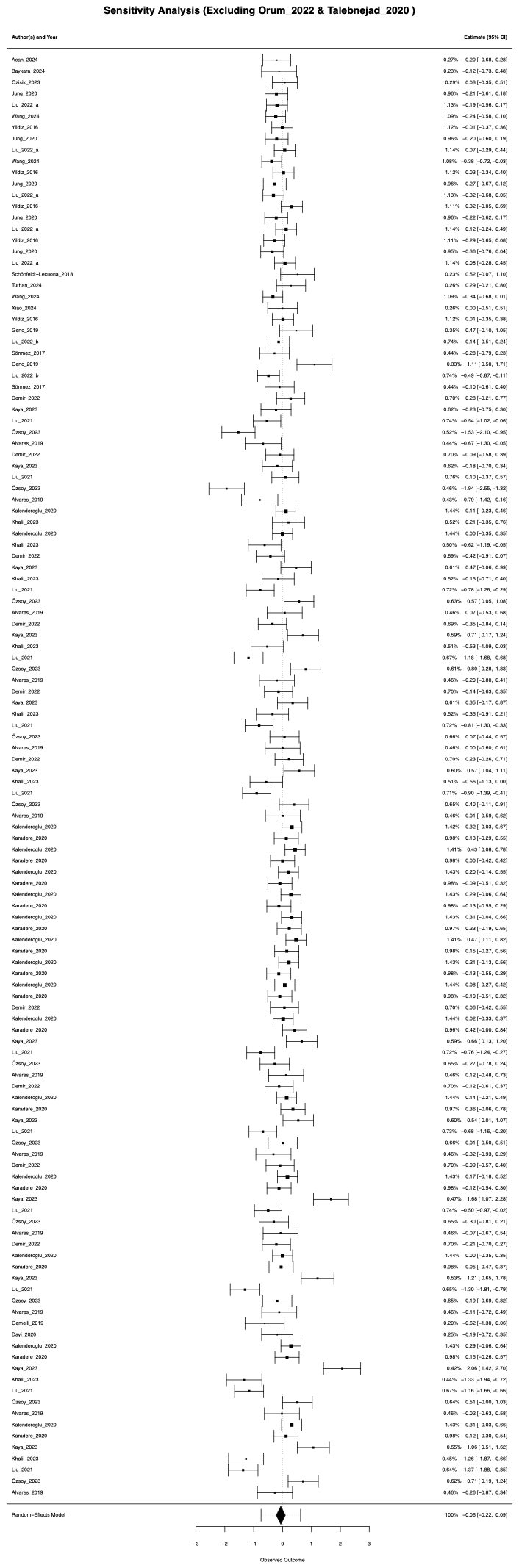

Funnel Plots:

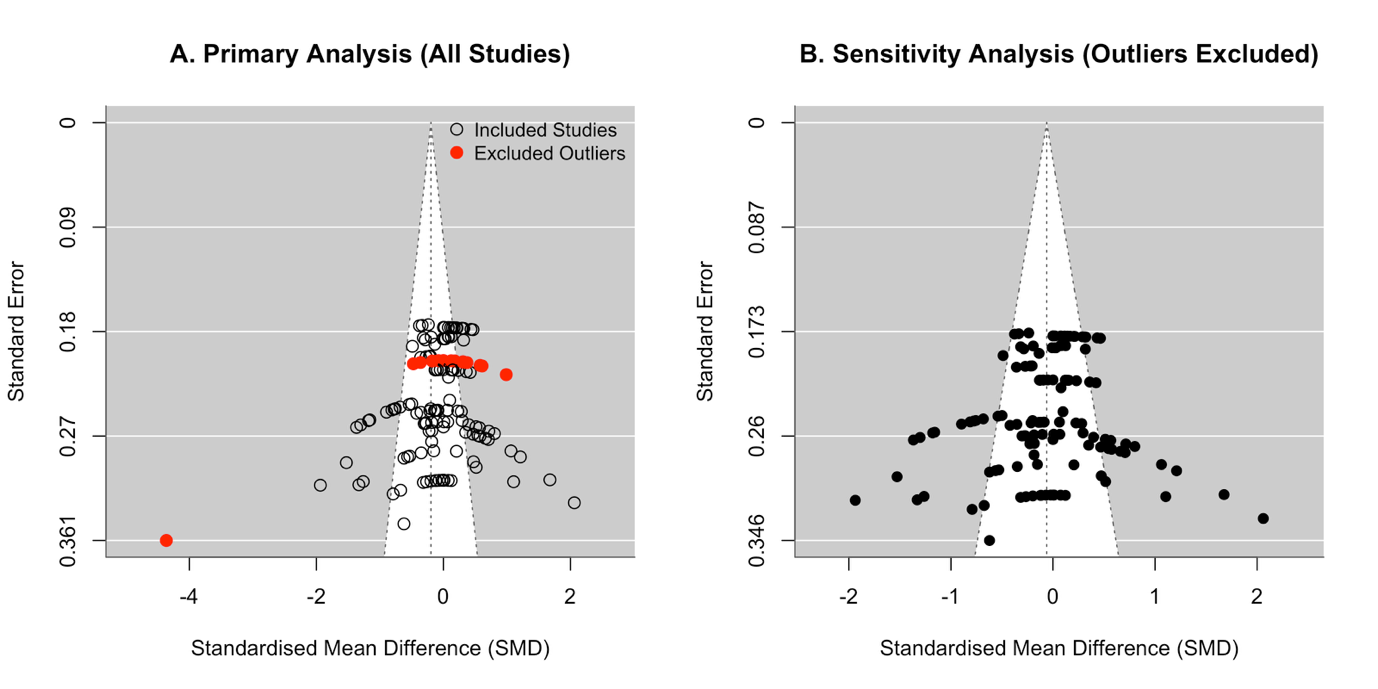

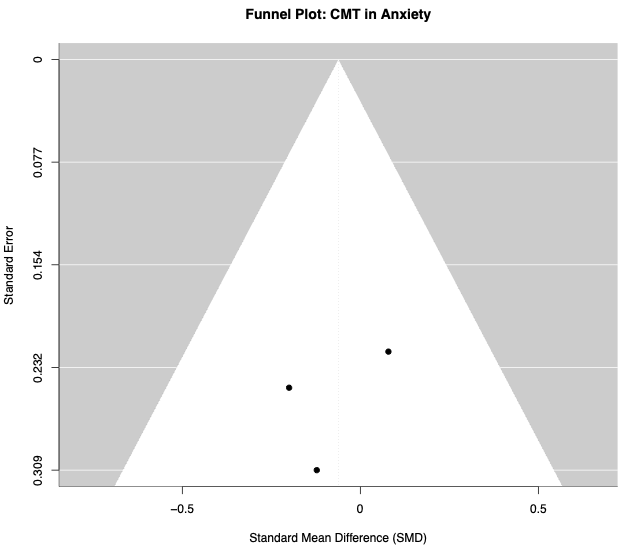

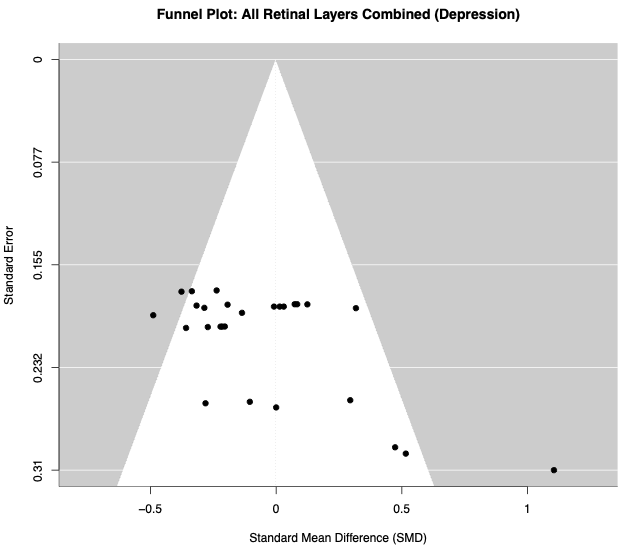

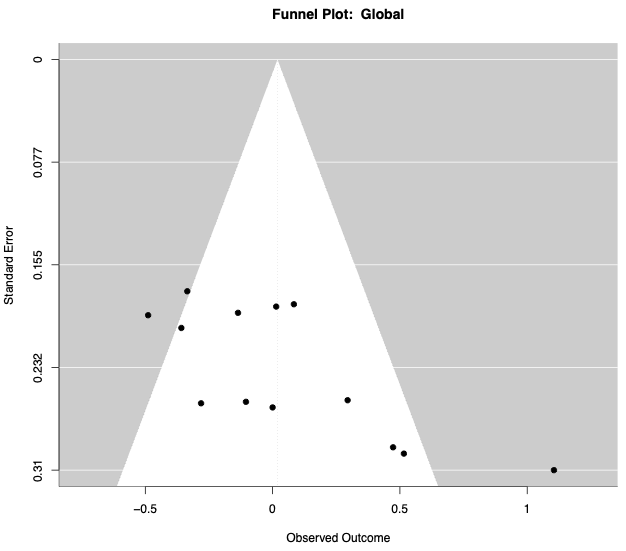

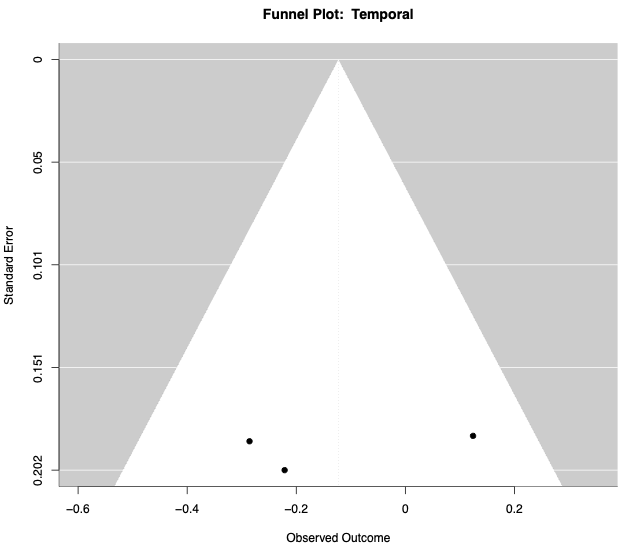

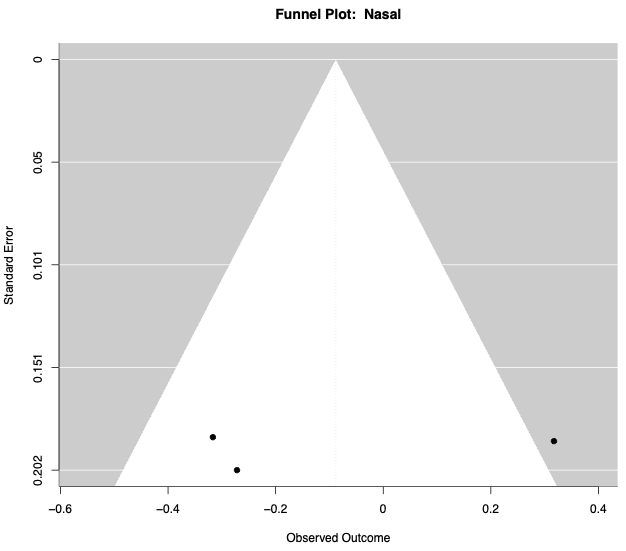

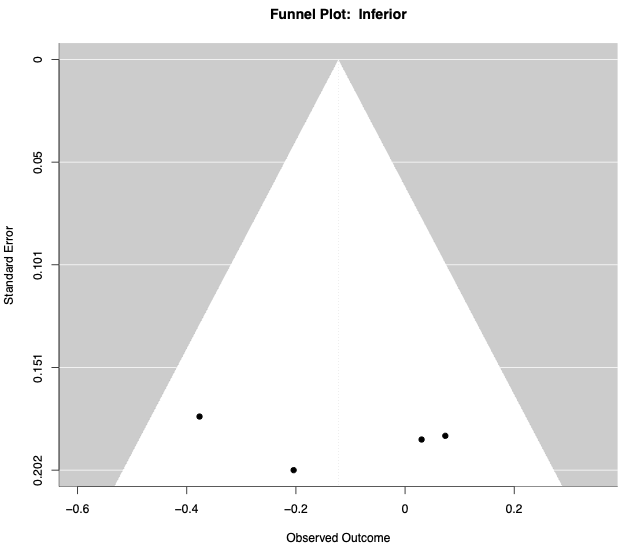

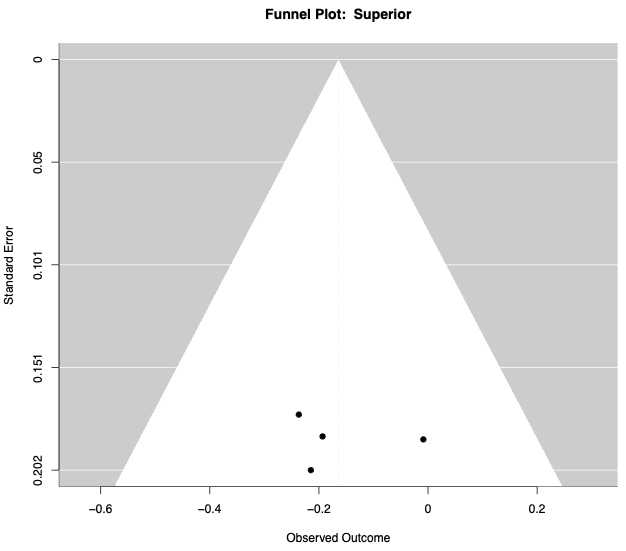

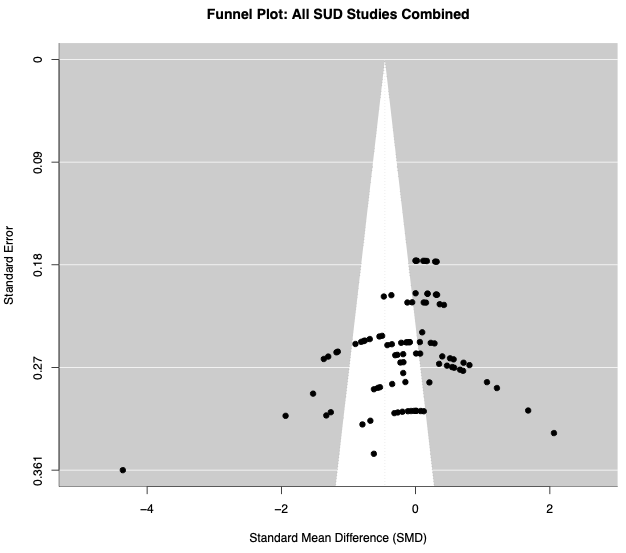

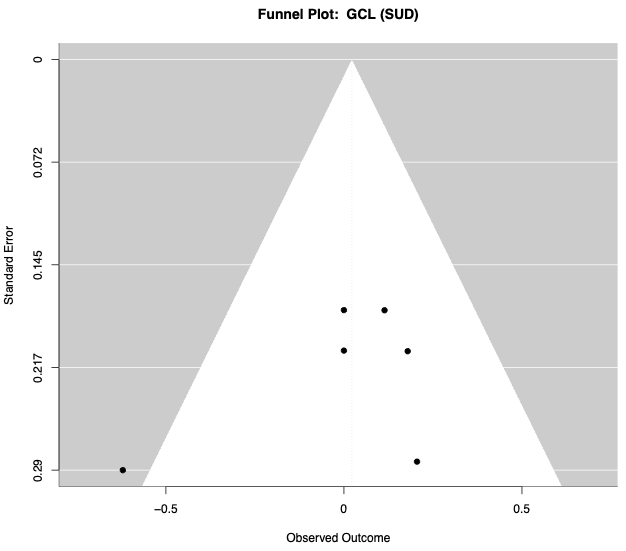

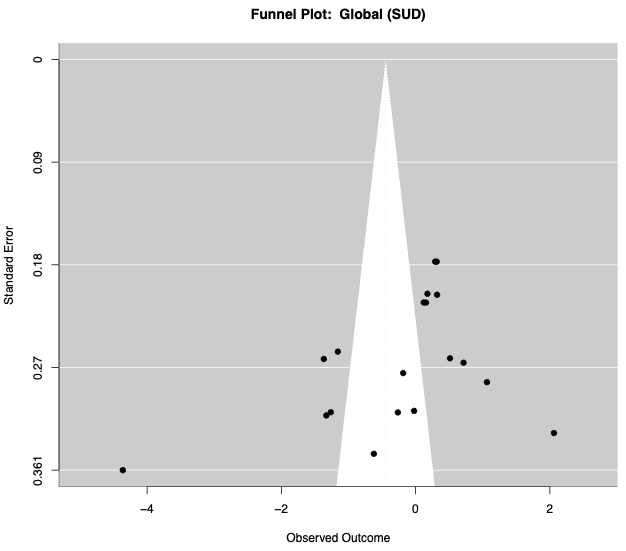

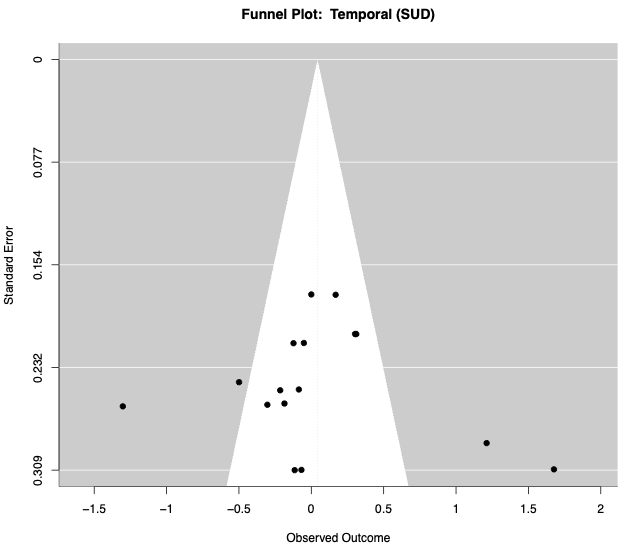

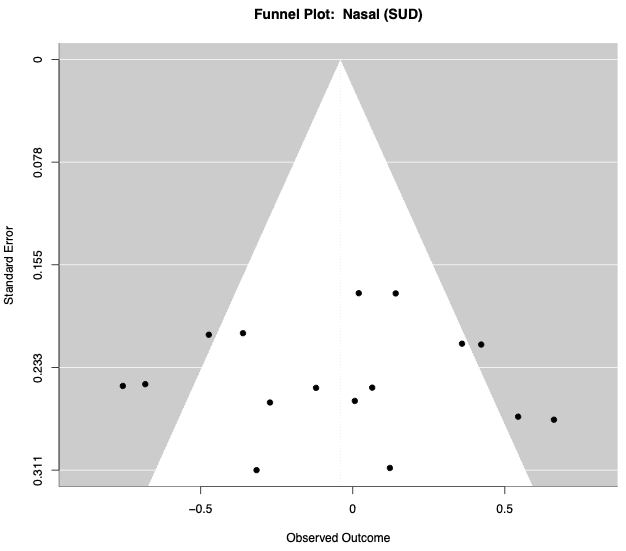

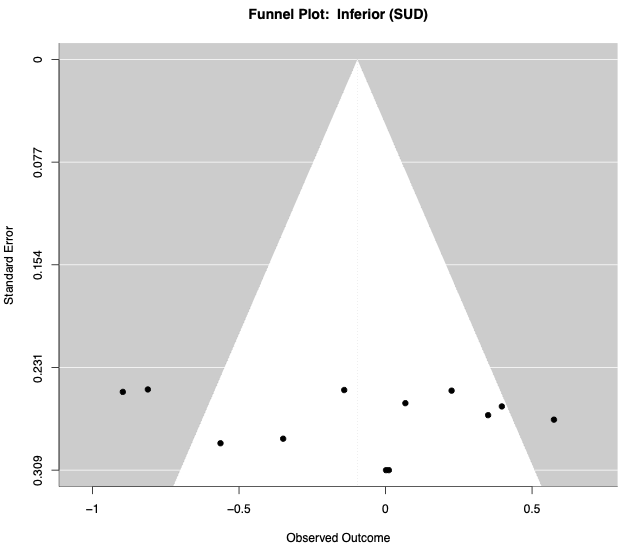

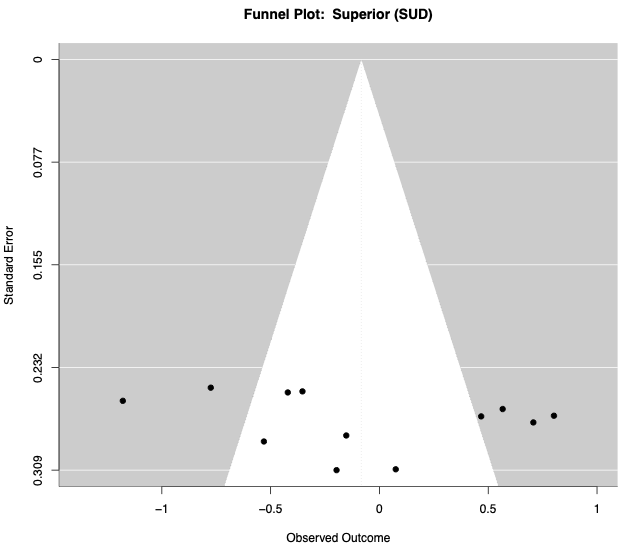

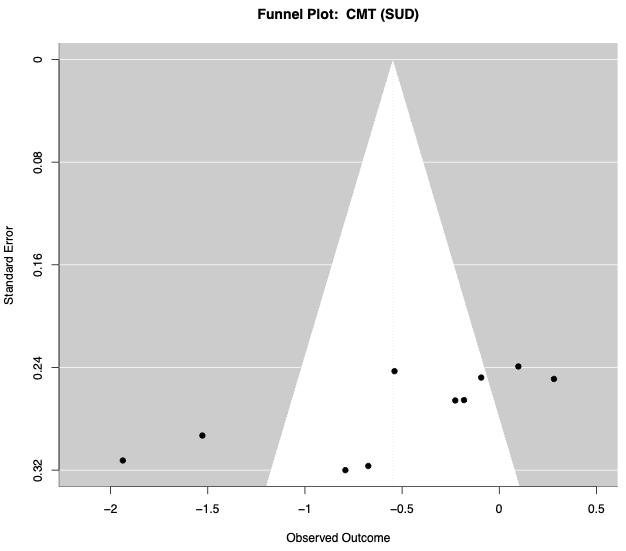
